# A Systems Biology Model Linking Peripheral NIMETOX Biomarkers to Multilevel Brain Structural and Functional Alterations Across Major Depression and Schizophrenia

**DOI:** 10.64898/2026.09.09.26362608

**Authors:** Hongzhou Wu, Chenghui Yang, Ying He, Xianfeng Qu, Abbas F Almulla, Yingqiang Zhang, Jinming Xiao, Chengxiao Yang, Drozdstoj Stoyanov, Bharat B Biswal, Benjamin Klugah-Brown, Andre F. Carvalho, Licia Pacheco Luna, Stefania Ferraro, Yuting Wang, Elijah Agoalikum, Michael Maes

## Abstract

Peripheral lipid and antioxidant-related biomarkers may help contextualize brain abnormalities across psychiatric disorders, but their relationships with brain structure, intrinsic activity, and dimensional symptoms remain incompletely characterized. In this cross-sectional study, 93 adults were recruited: 38 with major depressive disorder, 29 with schizophrenia, and 26 healthy controls. Eighty-seven participants met resting-state functional MRI quality-control criteria. Assessments included structural and functional MRI, serum lipids, albumin, apolipoprotein A-I, childhood maltreatment, psychological resilience, and clinical symptoms. Exploratory analyses combined regional imaging measures, within-network connectivity, statistical mediation, and partial least squares structural equation modeling. Group-related functional abnormalities involved parietal, sensorimotor, cingulate, insular, and temporal regions. Peripheral lipid and antioxidant-related measures were associated with regional gray matter measures and intrinsic activity. Childhood maltreatment and resilience were associated with mood-related symptom burden, whereas lower connectivity and nodal strength within the selected 11-region network were associated with psychosis-related and overall illness severity. Exploratory structural equation models accounted for 66.4% of the variance in the mood-related composite and 25.3% of the variance in local intrinsic activity. These findings describe cross-sectional relationships among peripheral biological measures, regional brain characteristics, developmental adversity, and psychopathology. They provide a framework for further investigation rather than evidence of a causal blood-to-brain sequence. Independent replication with prospectively defined measures and longitudinal assessments is needed.

## Introduction

Contemporary psychiatry faces major limitations in the diagnostic validity of conventional operational classifications, which remain predominantly based on clinical phenomenology rather than objectively measured biological mechanisms [1, 2]. Unlike other medical disciplines, psychiatric research also remains fragmented across clinical, molecular, and neuroimaging levels of explanation [3]. The nomothetic networks psychiatry framework was developed to overcome this fragmentation by integrating clinical phenotypes with peripheral biomarkers and brain-imaging measures within a bottom-up, data-driven systems-biology approach [4, 5]. Such multilevel integration may identify biologically meaningful pathways that cut across conventional diagnostic boundaries and underpin dimensional psychopathology. In this context, integrating peripheral biological processes with structural and functional brain alterations may provide a mechanistic framework linking systemic biological dysregulation to the neural substrates of major psychiatric disorders [3].

Although diagnostically distinct, major depressive disorder (MDD) and schizophrenia (SCZ) exhibit partially overlapping patterns of functional brain abnormalities, alongside disorder-specific alterations [6–9]. This overlap supports a transdiagnostic view in which dimensional symptoms arise from disruptions in large-scale systems rather than from abnormalities confined to isolated brain regions [8, 10, 11]. Resting-state fMRI provides complementary measures of low-frequency fluctuations (fALFF) and regional homogeneity (ReHo) to index local intrinsic activity, whereas functional connectivity (FC) and degree centrality quantify inter-regional communication and nodal integration within broader brain networks [12–15].

Alongside brain abnormalities, peripheral biological abnormalities may also contribute to the shared and disorder-specific pathophysiology underlying these dimensional symptoms. Increasing evidence indicates that patients with MDD and SCZ exhibit abnormalities in neuroimmune, metabolic, and oxidative stress (NIMETOX) signaling [16–18]. Lower albumin and apolipoprotein A1 (ApoA1) levels may represent convergent markers of these processes, as both proteins contribute substantially to antioxidant defenses and their reduction reflects a negative acute-phase response associated with inflammatory dysregulation [16]. ApoA1, the principal protein component of high-density lipoprotein (HDL) particles, participates in reverse cholesterol transport and has antioxidant and anti-inflammatory functions, whereas albumin contributes substantially to extracellular antioxidant buffering and ligand transport [19]. Atherogenic lipid indices and related lipid markers, including the atherogenic index of plasma (AIP), low-density lipoprotein cholesterol (LDL-C), very-low-density lipoprotein cholesterol (VLDL-C), and free cholesterol (F-C), have been implicated in major depressive disorder and bipolar disorder, particularly when comorbid with tobacco use disorder, as well as in schizophrenia, where they are associated with cardiovascular and metabolic risk, cognitive impairment, and adverse clinical outcomes [20–23]. Dyslipidemia may affect brain integrity through endothelial dysfunction, blood-brain barrier disruption, oxidative stress, and microglia-mediated neuroinflammatory pathways, potentially contributing to broader structural and functional brain alterations [24, 25]. In addition, network-neuroscience evidence suggests that highly connected hub regions support long-range communication and impose substantial metabolic and wiring costs, which may make them generally vulnerable to pathological perturbations [26, 27]. In the context of NIMETOX dysregulation, this vulnerability may help explain how systemic metabolic, oxidative, and inflammatory stress is linked to distributed structural and functional brain alterations [28, 29].

Furthermore, adverse childhood experiences (ACEs) are associated with distributed structural and functional alterations in the brain and with neurodevelopmental and immunometabolic pathways [30]. Resilience, by contrast, may buffer the association between childhood trauma and depressive symptomatology; meta-analytic and mediation evidence indicate that greater adversity is related to lower resilience and more severe depression [31, 32].

However, peripheral NIMETOX biomarkers, structural and functional brain measures, resilience-related factors, and clinical symptoms have often been examined either in isolation or in limited combinations, leaving their joint relationships insufficiently characterized. It therefore remains unclear whether these domains constitute a coherent blood–brain systems biology pathway linking peripheral biology to multilevel brain dysfunction and transdiagnostic clinical phenotypes, and whether such an integrated framework provides greater explanatory power than neuroimaging alone. We addressed this gap with a multilevel systems biology framework integrating peripheral antioxidant and lipid-related biomarkers, regional GMV, local intrinsic activity (fALFF/ReHo), functional connectivity (FC) and degree centrality (DC), ACE exposure, resilience, and dimensional clinical phenotypes across MDD, SCZ, and healthy controls. The imaging analyses began with voxel-wise analyses of fALFF and ReHo to identify regions showing group differences in local intrinsic activity. Regions of interest (ROIs) centered on the peaks of these significant functional clusters were then used to extract regional GMV and characterize interregional functional connectivity. Within this functionally defined spatial framework, mediation analyses examined two hypothesized indirect associations: peripheral biomarkers → GMV → local intrinsic activity, and GMV → local intrinsic activity → clinical symptoms. These associations were further integrated using exploratory partial least squares structural equation modeling (PLS-SEM). Functional connectivity and degree centrality within the selected ROI network were examined to characterize interregional coupling and its associations with dimensional clinical burden. Finally, orthogonal partial least squares discriminant analysis (OPLS-DA) and random forest analyses were used to evaluate the contributions of multimodal features to diagnostic group discrimination and dimensional prediction of illness burden.

Based on previous transdiagnostic neuroimaging evidence implicating parietal, sensorimotor, and cingulate regions in both MDD and SCZ, we hypothesized that the two disorders would show alterations in local intrinsic activity within these regions [33–35]. That a less favorable peripheral NIMETOX profile would be associated with lower GMV and weaker intrinsic activity; that reduced local and network-level function would track greater dimensional symptom burden; that ACE-related risk would be partly expressed through lower resilience; and that multimodal feature sets would outperform imaging-only models. This system’s biological framework aims to characterize interactions among peripheral biological processes, brain alterations, psychological factors, and clinical dimensions rather than define diagnostic classifiers. Accordingly, diagnostic groups were used as reference categories, while the primary objective was to identify continuous biological–brain–symptom relationships across the spectrum of mood and psychotic symptom severity.

## Materials and Methods

### Participants

We recruited 93 participants aged 18-65 years, including 38 patients with major depressive disorder (MDD), 29 patients with schizophrenia (SCZ), and 26 healthy controls (HCs). Patients with MDD and SCZ were recruited from the Mental Health Center of Sichuan Provincial People’s Hospital. HCs were recruited from family members of patients, hospital staff, and students at the University of Electronic Science and Technology of China. Diagnoses of MDD and SCZ were established by a senior psychiatrist according to DSM-5 criteria, using a structured clinical interview for DSM-5 psychiatric disorders [36].

Exclusion criteria included: age <18 or >65 years; standard MRI contraindications, including metallic implants or other MRI-incompatible devices; current or lifetime comorbid psychiatric disorders other than the primary diagnosis, including bipolar disorder, schizoaffective disorder, and substance use disorders other than nicotine dependence; autism spectrum disorder, borderline and antisocial personality disorders, or intellectual disability; neurological disorders, including stroke, epilepsy, brain tumor, Parkinson’s disease, Alzheimer’s disease, and multiple sclerosis; and major medical conditions likely to affect immune or metabolic function, including autoimmune diseases, psoriasis, systemic lupus erythematosus, inflammatory bowel disease, rheumatoid arthritis, type 1 diabetes mellitus, chronic obstructive pulmonary disease, and cancer. Additional exclusion criteria included pregnancy or breastfeeding; recent allergic or infectious conditions likely to affect inflammatory or immune-related biomarkers; current treatment with immunosuppressive or immunomodulatory agents; therapeutic use of antioxidants or omega-3 supplements within the previous 3 months; major surgery within the previous 3 months; frequent analgesic use; history of traumatic brain injury with loss of consciousness; and clinically relevant sleep disorders, including sleep apnea–hypopnea syndrome. Use of antipsychotic, antidepressant, or other psychotropic medication, as well as smoking behavior, was recorded for all participants and included as covariates in subsequent analyses.

We performed a *priori* power analysis using G*Power version 3.1.9.4, based on the most demanding structural equation in the primary statistical analysis (partial least squares structural equation modelling, PLS-SEM), with up to five explanatory variables predicting the endogenous outcomes. Assuming an effect size of 0.176, corresponding to approximately 15% explained variance, five predictors, α = 0.05, and statistical power of 0.80, the minimum required sample size was 79 participants.

The Ethics Committee of the University of Electronic Science and Technology of China approved this study (approval number: 30850). All participants provided written informed consent in accordance with the Declaration of Helsinki.

### Clinical Measurements

A trained senior psychiatrist conducted a clinical interview and administered the structured Mini-International Neuropsychiatric Interview (M.I.N.I. 6.0) [37, 38], the 21-item Hamilton Depression Rating Scale (HAMD-21) [39], and the Positive and Negative Syndrome Scale (PANSS) [40]. Additionally, all participants completed self-report scales to measure severity of depression and anxiety, namely the Beck Depression Inventory-II (BDI-II) and the State-Trait Anxiety Inventory (STAI; state version), respectively [29,30]. ACEs were assessed using the Childhood Trauma Questionnaire-Short Form (CTQ-SF) [43]. We used the total CTQ-SF score to reflect ACE load in our patients and controls. Resilience was assessed using the Connor-Davidson Resilience Scale [44]. Based on these clinical assessments, we constructed composite scores reflecting resilience, overall severity of mood disorders (OSOM), overall severity of psychosis (OSOP), and overall severity of illness (OSOI). We conceptualized OSOP as a z-unit-based composite score (z-sum of positive symptoms + z-sum of negative symptoms), as assessed with the PANSS. We constructed the OSOM index as the first principal component extracted from complementary indicators: the total Beck Depression Inventory (BDI-II) score, the State-Trait Anxiety Inventory (STAI), and binary MDD diagnostic status. We adopted this approach because no single measure adequately captures the full continuum of mood-disorder severity. Whereas the BDI-II and STAI quantify dimensional self-reported depressive and anxiety symptoms, the MDD diagnosis provides an independent clinician-established assessment based on a structured diagnostic interview. Consequently, the first principal component integrates subjective symptom severity with clinician-based diagnostic information into a single continuous latent variable representing overall mood-disorder severity. This continuous systems-level construct was considered more appropriate for mediation analyses and PLS-SEM than a binary diagnostic outcome. Before extraction, we evaluated sampling adequacy and unidimensionality using the Kaiser–Meyer–Olkin (KMO) statistic (KMO > 0.6), the proportion of explained variance (>50%), and item loadings on the first principal component (>0.70). The OSOM construct demonstrated adequate psychometric properties, with a Kaiser–Meyer–Olkin (KMO) measure of sampling adequacy of 0.636, the first principal component explaining 82.81% of the total variance, and all indicators exhibiting strong loadings on the latent construct (all loadings > 0.873). We conceptualized resilience as the first principal component (PC) extracted from the resilience scale items. Four items (items 2, 3, 9, and 20) failed to meet the loading criterion and were therefore excluded. We computed the final resilience score as the PC1 extracted from the remaining 21 items, denoted as PC-RISC. Suicidal ideation (SI) severity and lifetime suicidal behaviors (SB) were assessed using the Columbia-Suicide Severity Rating Scale, and current and lifetime SI and suicide attempts (SA) were computed as previously described [18]. The present study used current SI and total SB, the latter computed as a z-unit composite of current SI and lifetime SB. Body mass index (BMI) was calculated as body weight in kilograms divided by height in meters squared (kg/m^2^).

### Blood Collection and Processing

From 07:00 to 08:00 a.m., we collected a 10 mL fasting venous blood sample in serum tubes using disposable syringes. We centrifuged the samples at 3500 rpm for 10 minutes. We meticulously aliquoted the serum into Eppendorf tubes and stored it at −80 °C until subsequent analysis. The measurements of albumin, triglycerides (TG), total cholesterol (T-C), F-C, LDL-C, HDL-C, VLDL-C, and ApoA1 are listed in the Electronic Supplementary File (ESF), Table 1. Based on these values, we computed two different z-unit-based composite scores: a) z albumin + z ApoA1, two negative acute phase reactants with anti-inflammatory and antioxidant capacity (labelled as NAPR, negative acute phase response); and b) z TG – z HDL-C, which reflects the atherogenic index of plasma [23].

### MRI Data Acquisition

MRI data were acquired using a 3T MRI scanner (Siemens MAGNETOM Vida, Siemens Healthineers, Erlangen, Germany) equipped with a 64-channel head/neck coil at Sichuan Provincial People’s Hospital, Chengdu, China. High-resolution structural images were acquired using a 3D T1-weighted MPRAGE sequence with the following parameters: repetition time (TR) = 2300 ms, echo time (TE) = 2.26 ms, inversion time (TI) = 900 ms, flip angle = 12°, matrix size = 256 × 256, and slice thickness = 1 mm. Resting-state fMRI images were acquired using a 2D echo-planar imaging sequence while participants were instructed to keep their eyes open during the scan. The acquisition parameters were as follows: repetition time (TR) = 2000 ms, echo time (TE) = 29 ms, flip angle = 90°, matrix size = 80 × 80, slice thickness = 3 mm, spacing between slices = 3.75 mm, number of slices = 34, and phase-encoding direction = PA.

### MRI exclusion criteria

After preprocessing the resting-state fMRI data, we excluded participants with a mean framewise displacement > 0.2 mm or maximum head motion exceeding 1 mm translation / 1° rotation. Following these criteria, 5 MDD and 1 SCZ were excluded; the final 87 participants met these head motion criteria for the resting-state scans.

### R-St fMRI Data Preprocessing and Calculation of fALFF/ReHo

Image preprocessing was conducted using Data Processing & Analysis of Brain Imaging (DPABI, http://rfmri.org/DPABI). The first 10 volumes were discarded, followed by slice-timing correction, realignment, normalization to Montreal Neurological Institute space at 3-mm isotropic resolution, detrending, and nuisance regression using the 24-parameter Friston motion model and mean white matter and cerebrospinal fluid signals. Global signal regression was not performed. For fALFF analysis, the preprocessed but temporally unfiltered images were first smoothed with a 6-mm full-width-at-half-maximum Gaussian kernel, and fALFF was then computed as the ratio of the amplitude within the low-frequency range of 0.01–0.08 Hz to the amplitude across the entire detectable frequency range. For ReHo, smoothing was omitted prior to metric calculation, and the data were filtered between 0.01 and 0.10 Hz. FC preprocessing additionally included 6-mm smoothing and scrubbing of volumes with framewise displacement > 0.5 mm before nuisance regression, followed by 0.01-0.10 Hz filtering.

fALFF was calculated as the ratio of amplitude within 0.01-0.08 Hz to total spectral power and was z-standardized. ReHo was calculated using Kendall’s coefficient of concordance between each voxel and its 26 neighbors; the resulting maps were normalized by the global mean, smoothed with a 6-mm Gaussian kernel, and z-standardized. These normalized maps are referred to as fALFF and ReHo throughout.

### Voxel-wise Analysis of Covariance (ANCOVA)

After computing fALFF and ReHo maps for all participants, voxel-wise ANCOVAs were performed across the three groups, with age, sex, medication use, and BMI included as covariates. Because smoothed whole-brain maps exhibit strong spatial autocorrelation, multiple comparisons correction was conducted using Gaussian Random Field (GRF) theory. Gaussian random field (GRF) correction was applied to account for the spatial smoothness of the statistical maps and to control the cluster-level family-wise error rate. For the voxel-wise omnibus ANCOVA, statistical significance was determined using a cluster-forming voxel-level threshold of p < 0.001 and a cluster-level GRF-corrected threshold of p < 0.025. For subsequent pairwise post hoc comparisons, two-tailed tests were performed and corrected using the same GRF procedure.

### R-St Functional Connectivity and Nodal Centrality Calculation

To characterize interregional coupling among regions showing significant fALFF or ReHo abnormalities, seed-to-seed resting-state functional connectivity (FC) analysis was performed using Data Processing & Analysis for Brain Imaging (DPABI) [45]. To minimize potential bias arising from differences in cluster size, spherical regions of interest (ROIs) with a radius of 6 mm were centered on the peak MNI coordinates of the significant clusters identified in the fALFF and ReHo ANCOVA analyses. These ROIs were also used in the subsequent regional GMV analyses.

Following the preprocessing procedures described above, the mean BOLD time series was extracted from each ROI using DPABI. Pairwise Pearson correlation coefficients were calculated between the time series of all ROIs and subsequently transformed using Fisher’s *r*-to-*z* transformation to construct individual FC matrices. Weighted degree centrality, also referred to as nodal strength, was then calculated using the GRETNA toolbox [46]. For each node, weighted degree centrality was defined as the sum of the weights of all positive connections linked to that node. No sparsity threshold was applied; all positive-weight edges were retained, whereas non-positive edges were excluded.

### Gray Matter Volume Processing and Extraction

High-resolution T1-weighted images were processed using the CAT12 toolbox implemented in SPM12 to derive gray matter volume (GMV) maps. Images were segmented into gray matter, white matter, and cerebrospinal fluid, spatially normalized to Montreal Neurological Institute (MNI) space using DARTEL, and modulated using the corresponding Jacobian determinants to preserve regional tissue volume. The resulting modulated, normalized GM maps were resampled to an isotropic voxel size of 3 × 3 × 3 mm³ and smoothed with a 6-mm full-width-at-half-maximum Gaussian kernel.

To examine the structural correlates of the functional abnormalities, spherical regions of interest (ROIs) with a radius of 6 mm were centered on the peak MNI coordinates of the significant fALFF and ReHo clusters identified by the ANCOVA. For each participant, mean modulated GM values were extracted from the smoothed GM maps within each ROI. These regional GMV measures were entered into the mediation analyses, with total intracranial volume (TIV), estimated as the sum of gray matter, white matter, and cerebrospinal fluid volumes, included as a covariate to account for individual differences in head size.

### Statistical analysis

Conventional statistical analyses were performed using IBM SPSS Statistics version 30. Differences in categorical variables among the study groups were assessed using Pearson’s chi-square test. Continuous variables were compared using one-way analysis of variance (ANOVA), followed by Fisher’s least significant difference (LSD) post hoc tests for pairwise comparisons when the overall F-test was significant. Associations between continuous variables were evaluated using Pearson’s product-moment correlation coefficients. All statistical tests were two-tailed, and statistical significance was defined as p < 0.05.

### Systems biology and multivariate statistical analysis Mediation analysis

To examine relationships among blood factors, GMV, fALFF/ReHo, and clinical scales, we performed cross-sectional mediation analyses using the Multilevel Mediation and Moderation (M3) toolbox (https://github.com/canlab/MediationToolbox) [47, 48]. We tested two distinct mediation models. The first model examined whether GMV mediated the relationship between biomarkers and fALFF/ReHo. The second model examined whether fALFF/ReHo mediated the relationship between GMV and clinical scales. For both model types, linear regression models were used to estimate path a (X → M), path b (M → Y, controlling for X), the total association c (X → Y), and the direct association c′ (X → Y, controlling for M). The indirect association was quantified as a × b. Bootstrap-based inference was performed using 10,000 resamples of participants, and bias-corrected and accelerated (BCa) confidence intervals were calculated. All regression models included covariates of no interest to adjust for potential confounds (Age, Gender, and BMI). Statistical inference was focused on the indirect effect, and results were visualized with the toolbox’s path diagrams and histograms of the bootstrap distributions. Given the large number of mediation paths tested, we summarized all statistically significant mediation effects in a word-cloud representation (Python wordcloud package), where term size reflects the relative prominence of each variable across significant mediation pathways.

### Integrated biomarker analysis and systems biology modeling

We performed an integrated analysis of 56 imaging, lipid-related, oxidative stress, and ACE variables. Features were screened for excessive missingness, near-zero variance, and technical outliers. Variables with 10% or more missingness were excluded; for the remaining variables, missing values were imputed using k-nearest neighbors for the exploratory, univariate, and OPLS-DA analyses [49]. Skewed biochemical variables were log□-transformed for univariate testing, whereas continuous predictors were scaled to unit variance before OPLS-DA.

Principal component analysis was used to examine sample distribution and potential outliers. Between-group differences were assessed using Wilcoxon rank-sum tests, with p-values adjusted using the Benjamini–Hochberg false discovery rate procedure. Statistical significance was defined as FDR < 0.05. For strictly positive biochemical variables, fold changes >1.20 or <0.83 were additionally used to indicate increased or decreased levels, respectively, and rank-biserial correlations were calculated as effect-size estimates. Group discrimination was assessed using orthogonal partial least squares discriminant analysis (OPLS-DA) [50, 51]. Model performance was summarized using R^2^X, R^2^Y, and Q^2^ and validated by permutation testing, and nested cross-validation was used to assess out-of-sample classification performance and model generalizability. Variable importance in projection (VIP) scores were used to rank feature contributions to group separation.

### Regression analysis

Random Forest regression was used to predict OSOI and estimate feature importance [52]. Four feature sets were evaluated: imaging measures alone; imaging measures plus ACE variables; imaging measures plus blood biomarkers; and a 22-variable multimodal panel integrating imaging, blood biomarkers, and ACEs. Predictive performance was estimated using repeated nested cross-validation, with 5 inner, 5 outer, and 10 repeats, with preprocessing, imputation, feature selection, and hyperparameter tuning restricted to the training data. Inner folds were used for model optimization, and held-out outer folds were used exclusively for performance evaluation [53]. Model performance was quantified using R^2^, root-mean-square error, and mean absolute error. All analyses were conducted in R using RStudio (version 2026.01), primarily with the impute, ropls, randomForest, and caret packages. Model training, tuning, and resampling were implemented using the caret framework [54].

Multiple regression analyses were performed to identify independent predictors of the dimensional clinical scores. Final predictor selection combined manual and automated regression with overfitting control, ridge and best-subset regression, and evaluation of statistical performance and biological relevance within the NIMETOX framework. This approach minimized collinearity and overfitting while yielding parsimonious, biologically interpretable models. Selected predictors were subsequently entered simultaneously into manual conventional multiple regression models. The primary objective of the regression analyses was mechanistic explanation rather than clinical prediction. Standardized coefficients (β), t-values, exact p-values, and R^2^ were computed. Model assumptions were assessed using residual diagnostics, VIF and tolerance statistics, and White and modified Breusch–Pagan tests for heteroscedasticity. Model stability was examined using 1,000 bootstrap resamples; bootstrapped regression coefficients are reported only when they differ meaningfully from the coefficients obtained in the original multiple regression analysis.

### Partial Least Squares Structural Equation Modeling (PLS-SEM)

PLS-SEM was performed in SmartPLS 4 to integrate the principal pathways identified by the mediation analyses and to estimate their direct, indirect, and total associations. The structural model included antioxidant–anti-inflammatory status (NAPR), AIP, sensorimotor GMV, local intrinsic activity, ACE exposure, PC-RISC, OSOM, OSOP, and age. NAPR was specified as a formative composite comprising ApoA1 and albumin. Sensorimotor GMV was also specified formatively using mGMV04, mGMV07, mGMV09, mGMV10, and mGMV11. Local intrinsic activity was modeled reflectively using ReHo04, ReHo06, ReHo07, ReHo09, ReHo10, ReHo11, fALFF04, fALFF06, fALFF07, fALFF09, fALFF10, and fALFF11. ACEs, AIP, age, PC-RISC, OSOM, and OSOP were entered as single-indicator constructs. Age was modeled as an exogenous predictor of GMV and PC-RISC. PLS-SEM was selected because the model combined reflective and formative composites and focused on variance explanation and pathway estimation in a modest sample. PLS-SEM analyses were conducted only after the measurement model satisfied predefined quality criteria. Indicator reliability was considered adequate when all outer loadings exceeded 0.66 (p < 0.001). Construct reliability and convergent validity were confirmed by composite reliability > 0.80 and an average variance extracted (AVE) > 0.50. Reflective model specification was verified using Confirmatory Tetrad Analysis (CTA), whereas discriminant validity was established using the heterotrait–monotrait (HTMT) ratio. Model fit was considered acceptable when the standardized root mean square residual (SRMR) was < 0.10. Following confirmation of an adequate measurement model, the structural model was estimated using 5,000 bootstrap resamples to obtain standardized path coefficients, confidence intervals, and p-values, and to quantify direct, specific indirect, total indirect, and total effects. Predictive performance was evaluated using Stone–Geisser’s Q^2^ and Cross-Validated Predictive Ability Testing (CVPAT) with 10-fold cross-validation and out-of-sample prediction. A positive Q^2^ together with a statistically significant CVPAT result indicated that the PLS-SEM model exhibited meaningful predictive relevance and superior out-of-sample performance [55, 56].

## Results

### Sociodemographic, clinical, and biomarker data

Detailed descriptive statistics for the sociodemographic and clinical variables are provided in ESF, Table 2. Clinical assessments demonstrated distinct symptom profiles across diagnostic groups, with MDD and SCZ participants showing elevated psychopathological symptoms compared with HC participants. ESF, Table 3 shows the peripheral NIMETOX biomarkers.

### Regional intrinsic-activity abnormalities

Voxel-wise ANCOVA, controlling for age, sex, medication use, and BMI, revealed significant group differences in 7 fALFF clusters after GRF correction (**Figure 1A**). These clusters were predominantly distributed across bilateral parietal and sensorimotor regions, including the inferior and superior parietal lobules, postcentral gyri, paracentral lobule, middle cingulate cortex, and precuneus. The largest cluster was located in the left superior parietal lobule and postcentral gyrus, extending into the precuneus and inferior parietal lobule. **Table 1** provides detailed cluster sizes, peak coordinates, and F values.

**Figure 1.**
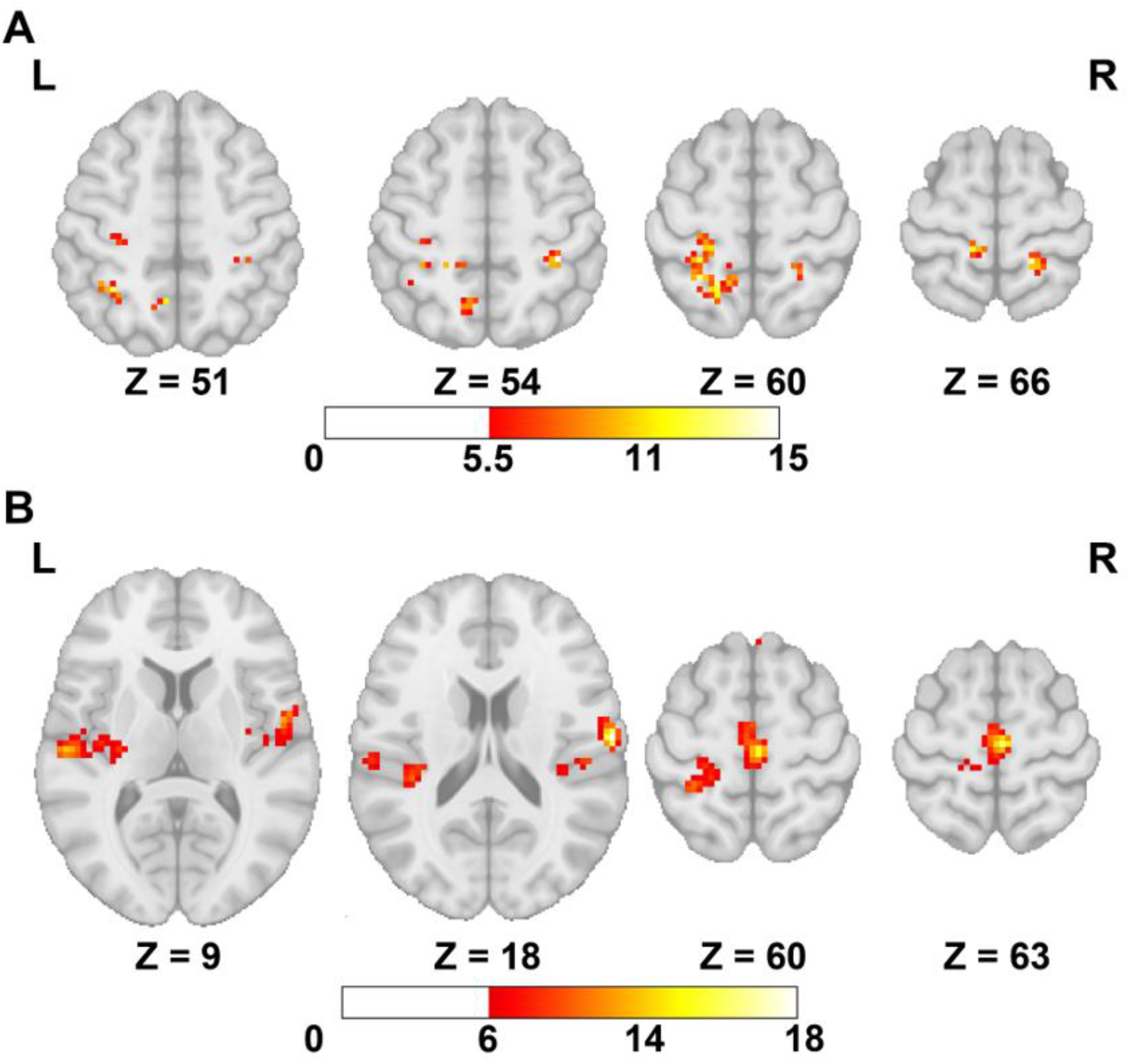
Covariance analysis (ANCOVA) of fALFF and ReHo across HC, MDD, and SCZ groups. (A) ANCOVA of fALFF. (B) ANCOVA of ReHo. The color bar represents the corresponding F-value.

The corresponding ReHo analysis identified four significant clusters (Figure 1B). The largest encompassed the bilateral supplementary motor area, the middle cingulate cortex, and the paracentral lobule and spatially overlapped with the medial fALFF abnormalities. Additional clusters were located in the right rolandic operculum–postcentral–insular region, the left postcentral/precentral cortex, and the left superior/transverse temporal–insular region. ReHo uniquely detected the latter.

Overall, fALFF and ReHo converged on abnormalities within bilateral sensorimotor and medial frontoparietal cortices, while ReHo additionally captured temporal and insular involvement. We then used the peak coordinates of these 11 clusters to define regions of interest for further analyses.

### Network disconnection and major mental disorders

The 11-node network showed higher mean connectivity in HCs (0.397 ± 0.128) than in MDD (0.269 ± 0.108) and SCZ (0.264 ± 0.104), with stronger interhemispheric coupling in HCs (**Figure 2A**). Edge-wise comparisons similarly revealed widespread connectivity reductions in both patient groups, particularly among connections involving ROI02, ROI07, ROI09, and ROI10 (Figure 2B). The ROI07–ROI09 connection showed the largest difference between HCs and the pooled patient group, whereas connections involving ROI01 were relatively preserved.

**Figure 2.**
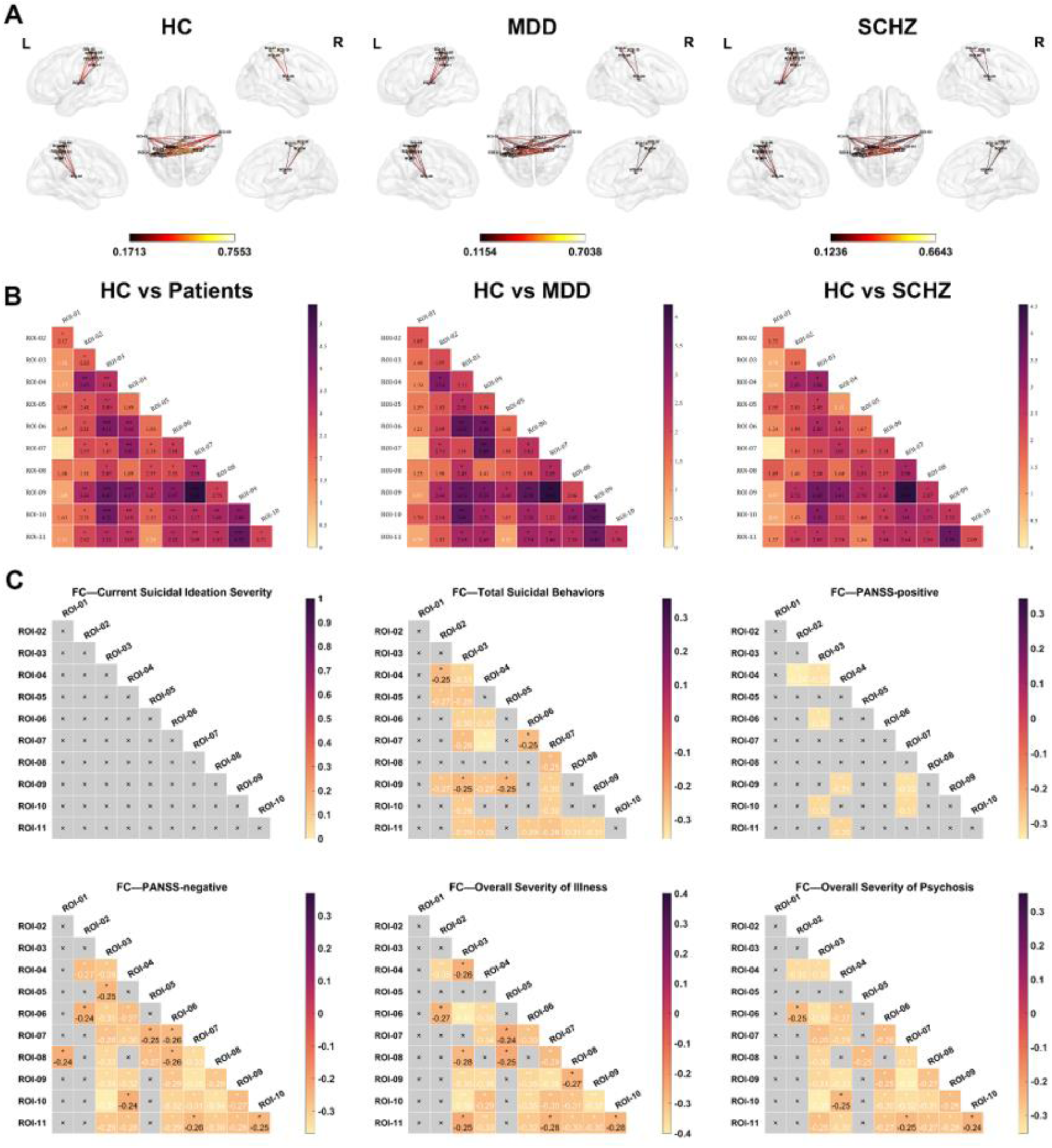
Resting-state functional connectivity within the 11-region disease-related network and its associations with clinical severity. (A) Mean within-network functional connectivity in the healthy control (HC), major depressive disorder (MDD), and schizophrenia (SCZ) groups. Nodes correspond to the 11 regions of interest identified by the fALFF and ReHo ANCOVAs; color bars indicate the mean Fisher-z-transformed connectivity strength within each group. L, left; R, right. (B) Lower-triangular heatmaps showing between-group comparisons of individual edges for HC versus the pooled patient group, HC versus MDD, and HC versus SCZ. Cell values represent the corresponding test statistics; positive values indicate stronger connectivity in HC. (C) Pearson correlation matrices between individual functional connections and Current Suicidal Ideation Severity, Total Suicidal Behaviors, PANSS-positive, PANSS-negative, Overall Severity of Illness, and Overall Severity of Psychosis. Cell values are Pearson correlation coefficients. Gray cells marked with “×” indicate non-significant associations at the threshold displayed. All *P* values after FDR correction. \**P* < 0.05; \*\**P* < 0.01; \*\*\**P* < 0.001.

Connectivity was also negatively associated with dimensional clinical burden (Figure 2C). Neither current suicidal ideation nor OSOM (not shown) was significantly associated with any individual edge, whereas total suicidal behaviors showed distributed negative correlations across the network (r = −0.25 to −0.36), strongest for ROI04–ROI07 (r = −0.36). These associations primarily involved cingulate, sensorimotor, opercular, supplementary motor, and temporal–insular regions. Psychosis-related severity showed progressively broader patterns of hypoconnectivity. PANSS-positive scores were associated with a relatively focal set of weaker connections (r = −0.30 to −0.34), whereas PANSS-negative scores showed more extensive associations (r = −0.24 to −0.37), strongest for ROI03–ROI10. OSOP was negatively associated with multiple edges (r = −0.24 to −0.35), while OSOI showed the broadest and strongest pattern (r = −0.24 to −0.40), peaking at ROI03–ROI06. Across clinical measures, all significant associations were negative, indicating that greater suicidal behavior and psychosis-related or overall illness severity were consistently associated with weaker network connectivity.

### Degree centrality and major mental disorders

Degree centrality was significantly lower in ROI02–ROI11 in both MDD and SCZ than in HCs, indicating widespread loss of nodal integration within the 11-region network (**Figure 3**). Lower degree centrality was also associated with greater clinical severity (ESF, Figure 1). Current suicidal ideation severity was negatively correlated with centrality in ROI03–ROI09 and ROI11 (r = −0.23 to −0.30), whereas Total suicidal behavior scores showed broader associations across ROI02–ROI11 (r = −0.23 to −0.33), strongest in ROI07 (r = −0.33). PANSS-positive scores were negatively associated with centrality in ROI02–ROI11 (r = −0.22 to −0.30), while PANSS-negative scores showed stronger associations across the same nodes (r = −0.27 to −0.36), peaking in ROI08. OSOP was similarly associated with lower centrality across ROI02–ROI11 (r = −0.26 to −0.34), and the strongest nodal association was observed between ROI06 and OSOI (r = −0.38). ROI01 was not significantly associated with any clinical measure. Together, these findings indicate that reduced nodal integration in the patient groups covaried with increasing suicidal and psychosis-related burden across a distributed network rather than a single hub.

**Figure 3.**
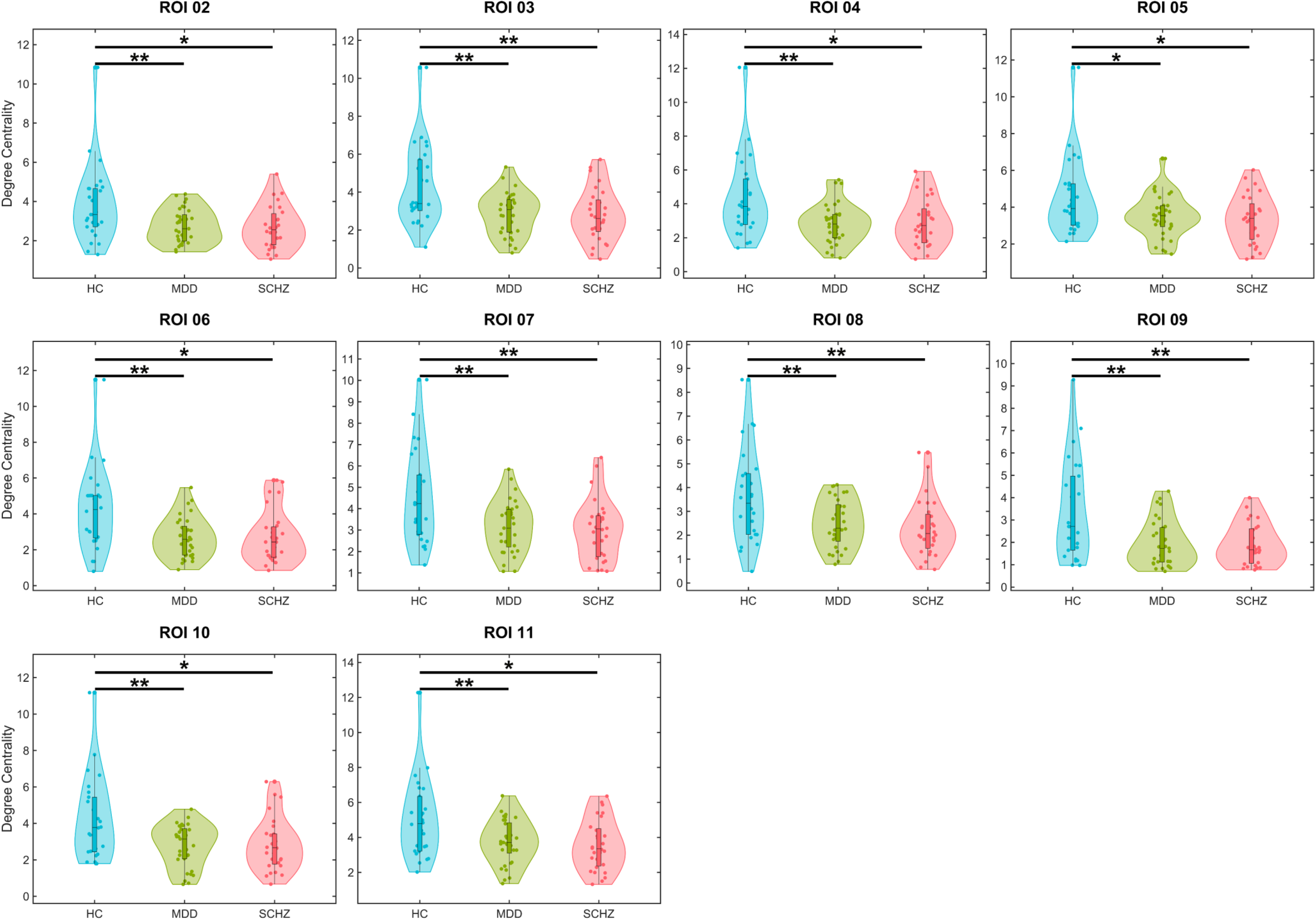
Group differences in nodal degree centrality and associations with dimensional clinical severity. Violin plots showing degree centrality in ROI02–ROI11 for the HC, MDD, and SCZ groups. Dots represent individual participants, and the internal summaries indicate the center and dispersion of each distribution. Group-comparison P values were corrected using the Benjamini–Hochberg false discovery rate procedure. All significant results passed the FDR correction. * p < 0.05, ** p<0.01, ***p<0.001.

### Multimodal discrimination and prediction

OPLS-DA was used to distinguish HCs from the pooled MDD and SCZ group (labeled as “mental disorders”) across MRI data, biomarkers, and ACEs (**Figure 4**). OPLS-DA revealed significant multivariate discrimination between healthy controls and patients with mental disorders (Figure 4 imaging-only). Permutation testing confirmed that the model was robust and not overfitted. In the full model integrating imaging measures, blood biomarkers, and ACE variables (Figure 4C), the variables with the highest VIP scores were fALFF11, fALFF02, ReHo09, ApoA1, and ReHo10. Nested cross-validation showed good out-of-sample discrimination, with an AUC of 0.824 and overall accuracy of 0.791. Sensitivity was relatively high (0.817), whereas specificity was somewhat lower (0.727). Balanced accuracy was 0.772, indicating reasonably balanced classification across controls and the pooled MDD/SCZ group. The F1-score of 0.847 indicates strong combined precision and sensitivity.

**Figure 4.**
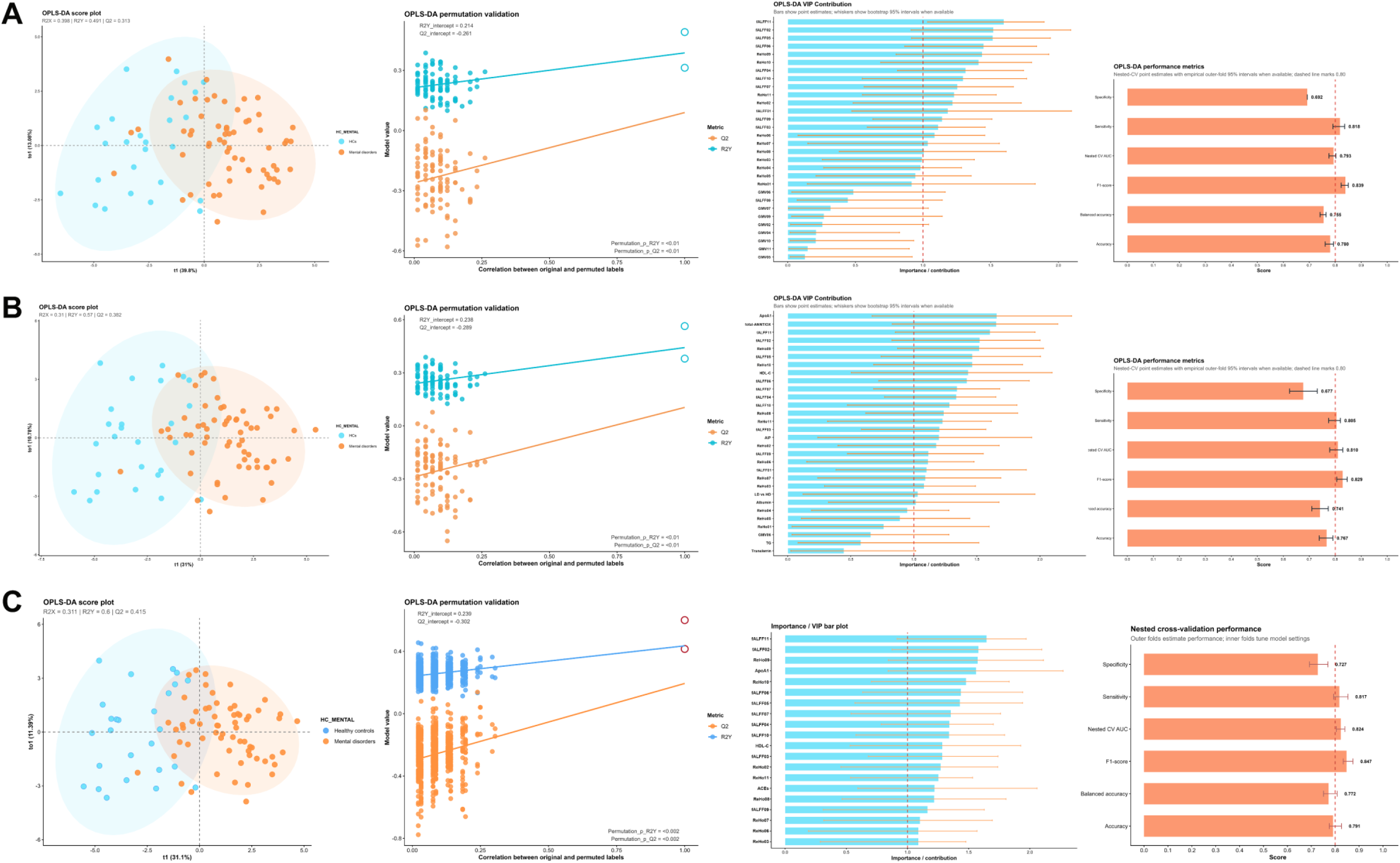
OPLS-DA discrimination between HCs and patients with mental disorders across different feature sets. For each model, we show the OPLS-DA plot, permutation validation, VIP contribution profile, and nested cross-validation performance metrics. (A) Brain imaging measures only (fALFF, ReHo, and GMV). (B) Brain imaging measures and blood biomarkers. (C) Brain imaging measures, blood biomarkers, and ACE variables.

### Explanatory systems biology analyses

**Figure 5** shows the results of random forest regression with OSOI as the dependent variable and MRI, ACE, and biomarker data as predictors. The most influential variables were NAPR, ACE exposure, fALFF11, ReHo09, and ReHo10 (Figure 5A). The model showed adequate predictive performance, with a strong correspondence between observed and predicted OSOI values (Figure 5B). Nested cross-validation (5 inner folds, 5 outer folds) with 1000 repeats produced out-of-sample performance estimates with a cross-validated R^2^ of 0.25, cross-validated RMSE of 0.836, and cross-validated MAE of 0.697, reflecting adequate model generalizability.

**Figure 5.**
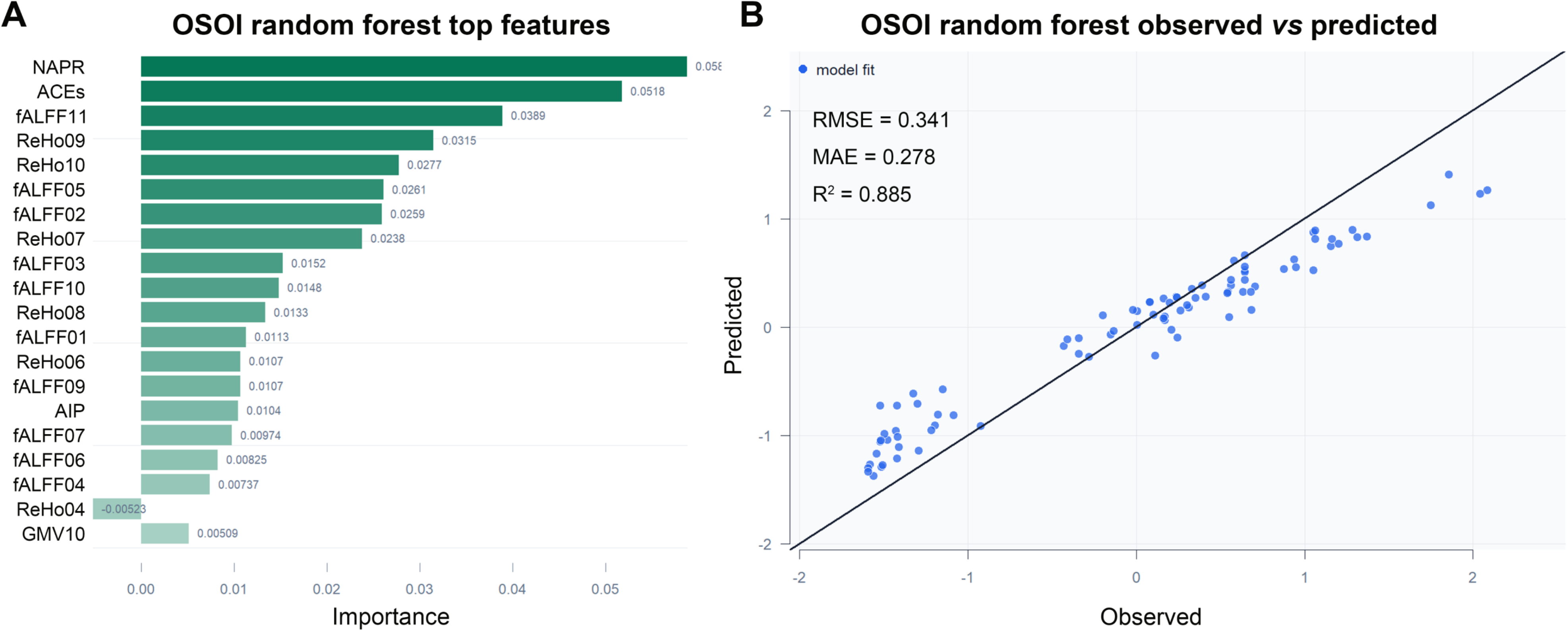
Random Forest regression model predicting the Overall Severity of Illness (OSOI). (A) Variable importance ranking identified NAPR (negative acute phase response), adverse childhood experiences (ACE), fALFF11, ReHo09, and ReHo10 as the top explanatory variables. (B) The right panel shows the correspondence between observed and Random Forest-predicted OSOI values in the complete dataset, demonstrating adequate predictive performance.

Based on the anatomical localization of the identified ROIs, z-unit–weighted composite scores were constructed for the sensorimotor, parietal, cingulate, and temporal regions (either GMV or fALFF/ReHo). These regional composite scores reduced dimensionality and multiple testing while capturing shared variance among anatomically and functionally related ROIs. They provide more stable, systems-level measures of regional brain structural and functional integrity than individual ROI measures. The sensorimotor composite comprised ROIs 4, 6, 7, 9, 10, and 11, encompassing postcentral, paracentral, and supplementary motor regions. The parietal composite comprised ROIs 1, 2, and 5 in the inferior/superior parietal lobules, while ROI3 represented the middle cingulate cortex and ROI8 the superior temporal gyrus. We additionally computed a sensorimotor Brain-Image composite as the z-unit–weighted sum of sensorimotor fALFF, ReHo, and GMV scores. This composite provides an integrated measure of sensorimotor structural integrity and intrinsic neural activity, thereby capturing their shared brain-imaging variance in a single systems-level index.

The correlation matrices in **Figure 6** indicate that the sensorimotor fALFF, ReHo, and GMV are associated with the clinical scores. Higher sensorimotor fALFF, ReHo, and the combined imaging score were associated with lower OSOI, OSOM, and OSOP scores. Sensorimotor GMV was associated with lower OSOI and OSOM, whereas its association with OSOP was not statistically significant at the threshold used in Figure 6. The imaging measures were positively associated with PC-RISC. ACE exposure shows the opposite pattern, being associated with poorer brain measures, greater illness severity, and lower resilience. Better sensorimotor structural and functional measures are positively associated with ApoA1 and NAPR and negatively associated with AIP. ApoA1 and NAPR are strongly correlated and both inversely related to AIP. Overall, the correlations support an integrated systems-biology pattern in which a more favorable peripheral antioxidant/negative acute-phase and lipid profile is associated with greater sensorimotor brain structural–functional integrity, which in turn is associated with resilience and lower dimensional psychopathology.

**Figure 6.**
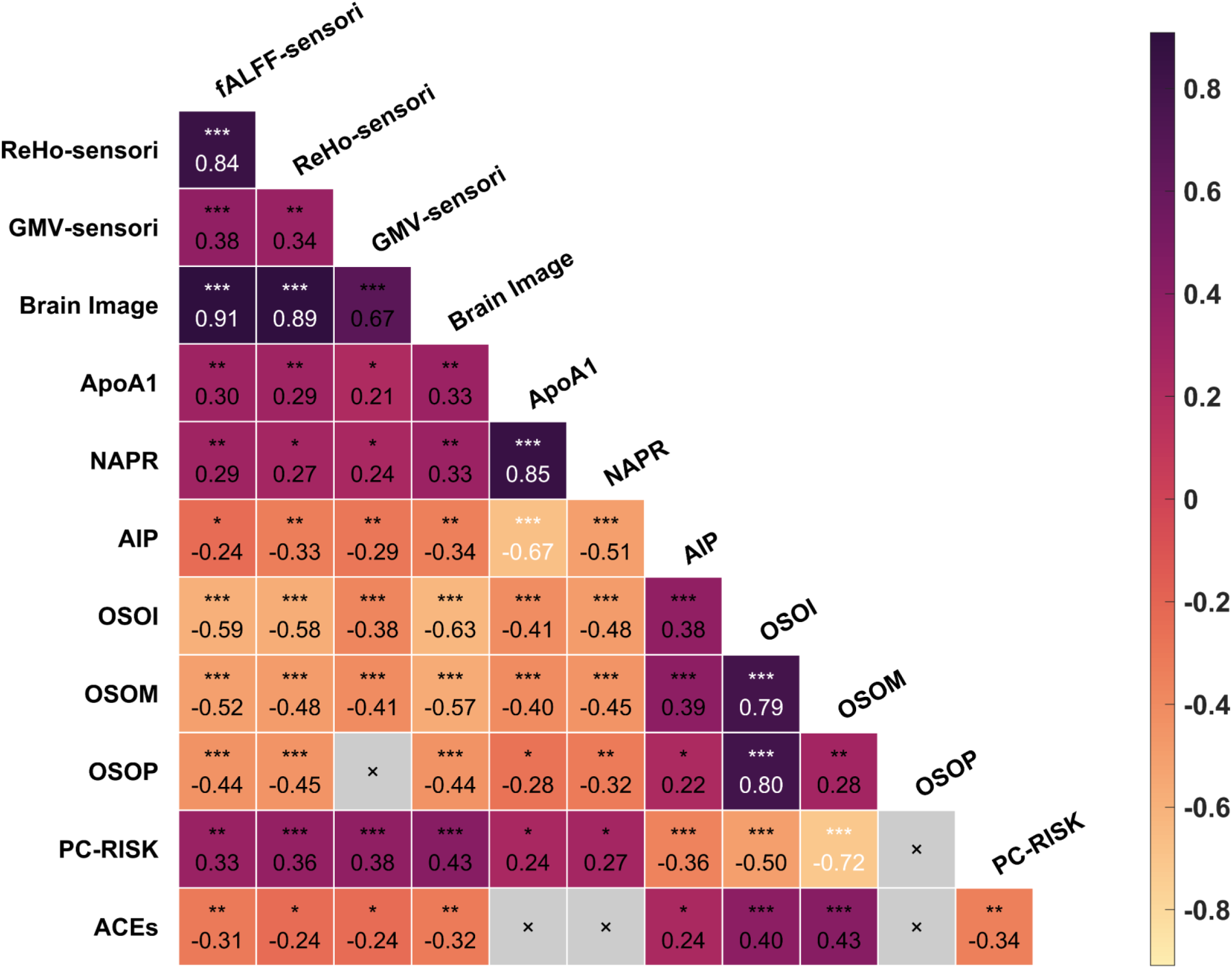
Correlation matrix between the sensorimotor fALFF, ReHo, and GMV values, NIMETOX biomarkers, and clinical scores. ApoA1: apolipoprotein A1; NAPR: negative acute phase response; AIP: atherogenic index of plasma; OSOI: overall severity of illness; OSOM: overall severity of mood disorders; OSOP: overall severity of psychosis; PC-RISC: psychological resilience; ACEs: adverse childhood experiences.

### Multiple regression

**Table 2** shows the results of multiple regression analyses with OSOI and PC-RISC as dependent variables. Regression #1 shows that 4 variables explained 61.8% of the variance in OSOI: PC-RISC, fALFF11, and NAPR (all inversely) and sexual abuse (positively). Regression #2 shows that ACEs and free cholesterol (both inversely) and ReHo010 and ReHo01 (both positively) explained 38.2% of the variance in PC-RISC. Regression #3 shows that 44.8% of the variance in the brain image composite was explained by 4 variables, namely age, sexual abuse, and VLDL-C (all three negatively) and NAPR (positively). Forced-entry adjustment for sex, BMI, age, and smoking showed no significant effects of these covariates, except for age in regression #3, and did not materially alter the significance of the primary predictors. Moreover, 1,000 bootstrap resamples confirmed the stability of the regression coefficients and model estimates.

### Mediation analysis: Biomarkers, GMV, and intrinsic activity

**Figure 7** presents two mediation systems-biology panels linking peripheral biology, brain structure/function, and psychopathology (A-B) and an integrated PLS-SEM (C). Figure 7A shows the links between peripheral biomarkers → GMV → intrinsic activity. The peripheral NIMETOX profile, dominated by AIP, VLDL-C, HDL-C, albumin, and ApoA1, was associated with regional GMV, which in turn was associated with intrinsic neuronal activity indexed mainly by ReHo and fALFF. The direct peripheral biology → intrinsic activity path indicates that peripheral biological alterations may also relate to brain function independently of GMV.

**Figure 7.**
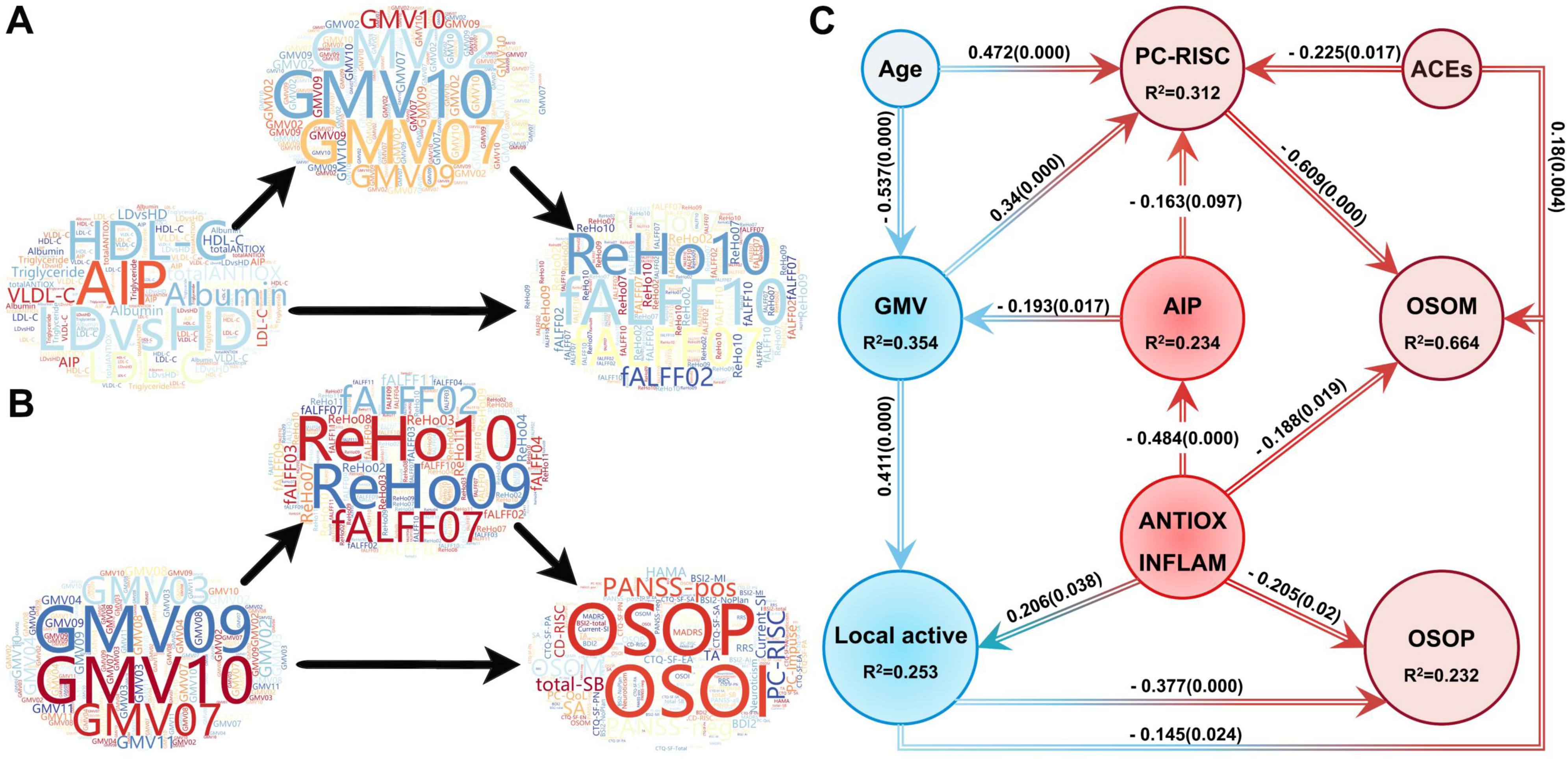
Integrated mediation summaries and partial least squares structural equation model linking peripheral biological status, regional brain structure and function, developmental adversity, resilience, and dimensional symptom burden. (A) Word-cloud summary of significant blood marker → GMV → local intrinsic activity mediation pathways. (B) Word-cloud summary of significant GMV → local intrinsic activity → clinical-outcome mediation pathways. In panels A and B, larger terms indicate greater relative prominence across the significant mediation pathways, and arrows indicate the modeled direction of the mediation sequence. (C) Results of PLS-SEM integrating antioxidant–anti-inflammatory status, AIP, sensorimotor GMV, local intrinsic activity, ACE exposure, PC-RISC, OSOM, and OSOP, with age included as an exogenous predictor. Antioxidant–anti-inflammatory status was formed by ApoA1 and albumin; the GMV composite comprised mGMV04, mGMV07, mGMV09, mGMV10, and mGMV11; and local intrinsic activity comprised ReHo04, ReHo06, ReHo07, ReHo09, ReHo10, ReHo11, fALFF04, fALFF06, fALFF07, fALFF09, fALFF10, and fALFF11. Values adjacent to structural paths are standardized path coefficients (β), with two-tailed p values from 5,000-sample bootstrap samples shown in parentheses; values displayed as 0.000 should be interpreted as p < 0.001. R^2^ values are shown within endogenous constructs.

Figure 7B shows the links from GMV → intrinsic activity → clinical phenome. Regional GMV, particularly involving GMV07, GMV09, and GMV10, predicts intrinsic activity characterized by ReHo and fALFF, which in turn predicts the dimensional clinical phenome (OSOP/OSOI and related symptom measures). The direct GMV → phenome pathway indicates additional structural associations with psychopathology not mediated by intrinsic activity. Thus, this figure supports a multilevel systems-biology cascade: peripheral biological imbalance → structural brain alterations → altered intrinsic neural activity → dimensional psychopathology, while retaining significant direct pathways between levels. Detailed pathway features are provided in ESF, Figures 2 and 3, and ESF, Tables 4 and 5.

### A systems biology PLS-SEM model

PLS-SEM was performed to investigate the direct and indirect relationships among ACEs, peripheral biomarkers, neuroimaging measures, and clinical phenotypes (**Figure 7C**). The local intrinsic activity construct (i.e., a factor extracted from the fALFF and ReHo ROIs 4, 6, 7, 9, 10, and 11) showed adequate reliability and acceptable convergent validity (Cronbach’s α = 0.922, rho_A = 0.927, composite reliability = 0.934, AVE = 0.545; loadings: 0.592–0.827 (all p < 0.001). Sensorimotor GMV was modeled as a formative construct comprising the GMV values of ROI04, ROI07, ROI09, ROI10, and ROI11, whereas all other variables were modeled as single-indicator constructs. Discriminant validity was established using the heterotrait–monotrait ratio (HTMT), with all HTMT values meeting the recommended threshold criteria. The saturated- and estimated-model SRMR values were 0.079 and 0.094, respectively. PLSpredict showed positive predictive relevance (Q^2^predict > 0), and PLS-SEM significantly outperformed the indicator-average benchmark (CVPAT, p = 0.001). Overall, the model shows good predictive relevance, with its principal strength being latent-construct modeling and structural-theory testing.

We found that 66.4% of the variance in OSOM was explained by the regression on PC-RISC, NAPR, and ACEs, whilst 23.2% of the variance in OSOP was explained by local intrinsic active and NAPR. Importantly, 31.2% of the variance in PC-RISC was explained by the regression on AIP (risk factor), GMV, and age (both protective). The model showed that 25.3% of the variance in local activity was explained by GMV and NAPR combined.

**Table 3** presents the total effects (direct + indirect effects) among peripheral biomarkers, brain structural–functional, resilience, and clinical constructs in the PLS-SEM model. Significant effects support integrated pathways linking NAPR and AIP with sensorimotor GMV and intrinsic functional activity, which are subsequently associated with the clinical phenome. The model also identifies an ACE–resilience–mood pathway, with resilience (Z-score of PC-RISC) showing a particularly strong relationship with OSOM. ESF, Table 6 and 7 show the total indirect effects and the specific indirect effects, respectively.

### Effects of drug status on brain imaging and biomarker data

Some patients were receiving antidepressants (n = 33), benzodiazepines (n = 36), mood stabilizers (n = 26), or antipsychotics (n = 49). Potential medication effects were examined using multivariate GLM analyses of the MRI measures (sensorimotor, parietal, insular, and temporal fALFF, ReHo, and GMV) and peripheral biomarkers, with medication-use status, age, sex, and BMI entered as explanatory variables. No significant multivariate effects on MRI or biomarker measures were observed for antidepressants (F = 0.77, df = 12/72, p = 0.674, and F = 0.57, df = 8/76, p = 0.796, respectively), benzodiazepines (F = 0.78, df = 12/72, p = 0.666; F = 0.62, df = 8/76, p = 0.760), mood stabilizers (F = 0.70, df = 12/72, p = 0.750; F = 1.06, df = 8/76, p = 0.401), or antipsychotics (F = 0.90, df = 12/72, p = 0.551; F = 1.56, df = 8/76, p = 0.150). Moreover, none of the corresponding univariate GLM effects were significant, indicating no detectable association between current medication-use status and the MRI or peripheral biomarker measures examined. In addition, these treatments did not have a significant effect on functional connectivity and degree centrality. Smoking behavior did not have a significant effect on any of the brain imaging data. GLM analyses with MDD and smoking as fixed factors, age, sex, and BMI as covariates (multivariate and test of between subject effects) showed no significant effects of smoking on the serum biomarker data (see ESF, Table 2).

## Discussion

In this study, we employed a hierarchical blood–brain systems biology approach integrating peripheral lipid-related and antioxidant-antiinflammatory defense markers with regional GMV, resting-state intrinsic brain activity, functional connectivity, developmental adversity, psychological resilience, and dimensional clinical phenotypes to delineate shared neurobiological mechanisms across MDD and schizophrenia. The findings converged on a parietal–sensorimotor–cingulo-opercular system and supported a multilevel framework in which peripheral NIMETOX imbalances were associated with regional structural integrity, local intrinsic neural activity, network organization, and clinical burden. Moreover, multivariate analysis showed that brain imaging, peripheral NIMETOX biomarkers, and early-life stressors provided complementary discriminatory information. Mediation analyses and PLS-SEM consistently linked a less favorable antioxidant–atherogenic profile to lower GMV and weaker intrinsic activity, whereas ACE exposure was associated with mood-related symptoms partly through reduced resilience. Moreover, childhood sexual abuse was independently associated with the sensorimotor brain-image composite, integrating GMV, fALFF, and ReHo abnormalities. Functional connectivity and degree-centrality analyses further showed that regional abnormalities were embedded within a distributed network whose reduced integration covaried with suicidality and psychosis severity. Together, these results support an integrated ACE-peripheral NIMETOX pathways–GMV alterations-brain functional dysregulation–dimensional clinical symptoms model.

### A shared parietal–sensorimotor–cingulo-opercular substrate across disorders

The fALFF and ReHo analyses identified convergent abnormalities in bilateral parietal and sensorimotor cortices, the paracentral lobule, middle cingulate cortex, supplementary motor area, and temporal–insular regions. Although psychiatric neuroimaging has traditionally emphasized prefrontal–limbic dysfunction, increasing evidence implicates sensorimotor and parietal systems in both MDD and schizophrenia [57, 58]. Rather than constituting a single functional system, these regions participate in partially overlapping parietal, sensorimotor, cingulate, and insular networks involved in sensorimotor integration, action preparation, and the processing of bodily and behaviorally salient information [59–61]. Convergent dysfunction across these interacting networks may therefore contribute to dimensional symptom burden across MDD and schizophrenia.

The convergence of fALFF and ReHo abnormalities strengthens the interpretation that these findings reflect a coherent disturbance of local neural dynamics rather than isolated regional effects. fALFF captures the relative amplitude of spontaneous low-frequency fluctuations, whereas ReHo reflects the local synchronization of spontaneous neural activity; their spatial convergence therefore indicates complementary alterations in oscillatory activity and regional coordination of intrinsic brain activity. ReHo additionally detected a superior temporal–insular region, suggesting that local synchronization may be particularly sensitive to auditory, interoceptive, and salience-related abnormalities relevant to schizophrenia and depression [62, 63]. Overall, these results define a regional substrate of altered intrinsic brain function that provides the basis for the subsequent integration with structural, network-level, peripheral biomarker, and clinical findings.

### Peripheral NIMETOX biomarkers, GMV, and intrinsic neural activity

The central mechanistic finding was the convergence of the mediation and PLS-SEM results on a peripheral NIMETOX-GMV-brain function pathway. Lowered anti-inflammatory and antioxidant defenses (due to a negative acute phase response) were associated with increased atherogenicity, while the latter was associated with lower GMV, and lower GMV was associated with weaker intrinsic activity. The integrated GMV–intrinsic activity index was predicted by NAPR, increased VLDL-C, and childhood sexual abuse, linking peripheral NIMETOX dysregulation and developmental adversity to brain dysfunction. These findings support regional structural integrity as a potential intermediate substrate linking peripheral NIMETOX processes to local intrinsic brain function.

ApoA1 is central to HDL-mediated reverse cholesterol transport and has antioxidant and anti-inflammatory properties, whereas albumin contributes substantially to circulating antioxidant buffering [19, 64]. AIP reflects the balance between triglycerides and HDL-related protection and is associated with an atherogenic lipid profile [20]. A shift toward lower antioxidant/anti-inflammatory protection and greater atherogenicity may influence the brain through vascular dysfunction, oxidative stress, disruption of the blood–brain barrier, and neuroimmune activation [16, 65]. These processes may compromise cellular, synaptic, and structural integrity, particularly in highly connected regions with substantial metabolic and information-processing demands. Within this framework, GMV appears to represent more than a parallel imaging correlate. Its association with both intrinsic activity and resilience positions regional structure as a potential intermediate substrate linking peripheral NIMETOX pathways to functional brain alterations and clinical variation.

The PLS-SEM also identified a parallel developmental pathway. Greater ACE exposure was associated with lower resilience and greater mood symptom severity, while resilience showed a robust association with mood symptom severity in the model. The significant ACEs – resilience – mood indirect (mediated) effect is consistent with evidence that resilience may attenuate the relationship between childhood adversity and depression [31, 32]. Importantly, GMV was also positively associated with resilience, suggesting that the peripheral biological and developmental pathways converged rather than operating independently. Regional brain integrity may therefore represent a point of convergence between developmental adversity, biological vulnerability and adaptive psychological capacity.

Interestingly, the pathways to mood- and psychosis-related symptoms were partially differentiated. Mood-related symptom severity was associated with ACE exposure, psychological resilience, peripheral NIMETOX defenses, and intrinsic activity, whereas dimensional psychosis symptoms showed a stronger association with intrinsic activity. This pattern suggests that developmental adversity and resilience may be especially relevant to affective symptom expression, while disruption of sensorimotor intrinsic dynamics may contribute more directly to psychosis-related symptoms.

### Network disintegration as a functional expression of regional vulnerability

The functional connectivity and degree-centrality analyses extended the regional findings from local neural activity to network organization. Both patient groups showed widespread reductions in connectivity and nodal integration within the 11-region network, particularly among parietal, postcentral, cingulate, opercular, temporal–insular, and supplementary motor regions. Thus, the regions implicated in the GMV and intrinsic-activity pathways were also embedded within a distributed network characterized by reduced interregional communication and topological integration.

The clinical correlations further indicated that network disruption varied continuously with symptom severity. All significant edge- and node-level associations were negative: greater cumulative suicidal behavior, positive and negative psychotic symptoms, and psychosis severity were associated with weaker connectivity and lower degree centrality. These findings support a distributed loss of functional integration rather than dysfunction confined to individual connections or hubs. This interpretation is consistent with dysconnectivity models of schizophrenia and with evidence that altered functional-network centrality involves the postcentral cortex, insula, and parietal association regions [66, 67].

Current suicidal ideation was associated with reduced nodal centrality, whereas cumulative suicidal behavior was associated with both hypoconnectivity and reduced centrality. This may suggest that current ideation reflects a more dynamic state, whereas cumulative suicidal behavior reflects more persistent and distributed network dysfunction. This interpretation is consistent with reports that the functional connectivity correlates of suicidal behavior are not equivalent to those of current ideation severity and may be distributed across the connectome rather than localized to a single region [68, 69].

Psychosis-related associations also followed a graded dimensional pattern. Positive symptoms were related to a comparatively restricted set of altered functional connections and nodes, whereas negative symptoms and broader psychosis or illness severity showed more widespread network-level associations. Previous work has similarly linked sensorimotor hypoconnectivity and reduced network centrality to negative symptoms and clinical severity in schizophrenia [67, 70].

### Multimodal evidence for the integrated blood-brain framework

The OPLS-DA and random forest analyses provided complementary multivariate support for the systems biology NIMETOX framework established in this study [16, 65]. Adding peripheral biomarkers and ACE measures complemented rather than replaced the imaging signal, with influential predictors spanning brain imaging, NIMETOX biomarkers, and developmental adversity. Importantly, the most informative imaging features involved the same sensorimotor–parietal regions identified by the voxel-wise, mediation, and network analyses, providing convergent evidence across analytical approaches.

Taken together, the classification and regression analyses provided convergent systems biology evidence linking peripheral NIMETOX dysregulation and developmental adversity to structural and functional brain alterations and dimensional psychopathology. The findings support a principal NIMETOX → GMV → local intrinsic activity → network disintegration → clinical symptoms (and diagnosis of “major mental disorders”) pathway, with ACE exposure and resilience forming parallel pathways particularly related to mood symptoms.

### Limitations and strengths

Several limitations should be considered. First, the cross-sectional design precludes temporal and causal inference, and the directionally specified mediation and PLS-SEM paths may reflect reciprocal relationships or residual confounding. Second, although the sample was adequately powered for the planned explanatory systems biology and regression analyses, it was recruited at a single center. Therefore, the proposed NIMETOX–brain model requires independent replication and external validation in cohorts from different countries, ethnicities, and cultural backgrounds to establish its generalizability. Third, peripheral antioxidant and anti-inflammatory homeostasis was represented by the major negative acute-phase reactants ApoA1 and albumin. Future explanatory systems biology studies should broaden this biomarker panel by incorporating measures of oxidative and nitrosative damage, fatty acid composition, metabolomics, lipidomics, neurotrophic factors, and additional immunometabolic pathways to achieve a more comprehensive characterization of the NIMETOX–brain axis. Fourth, the network was restricted to 11 regions selected from group-level intrinsic-activity differences in the same cohort. This data-dependent definition improves anatomical specificity but may inflate apparent network effects and does not capture the position of these regions within the whole-brain connectome. Independent ROI validation and whole-brain analyses are needed. The absence of significant effects of medication-use status (use versus non-use) indicates that the primary contrast in medication exposure did not influence the MRI or biomarker findings; consequently, subordinate effects related to dose and treatment duration were not further examined. However, the present results cannot exclude such effects within treated patients.

The study also has several strengths. Structural MRI, complementary measures of local intrinsic activity, functional connectivity, peripheral biological markers, developmental adversity, resilience, and dimensional symptoms were assessed within the same cohort. The analysis combined pathway-oriented methods with edge- and node-level network measures and multivariate prediction. Most importantly, the convergence of findings from mediation, PLS-SEM, connectivity, degree centrality, OPLS-DA, and random forest enabled the different analyses to inform a single biological framework rather than generate disconnected, modality-specific conclusions.

## Conclusions

This hypothesis-driven systems biology study identified a shared parietal–sensorimotor–cingulo-opercular dysfunction across MDD and SCZ. Mediation and PLS-SEM supported a hierarchical pathway linking an unfavorable antioxidant–atherogenic profile to reduced GMV and intrinsic activity, with network disintegration associated with suicidality and mood and psychosis severity. ACE exposure additionally contributed to affective symptoms partly through reduced resilience. Multivariate analyses confirmed complementary contributions of imaging, NIMETOX, and developmental factors. Collectively, the findings support a multilevel NIMETOX–brain framework describing associations among peripheral dysregulation, developmental adversity, brain dysfunction, and dimensional psychopathology, requiring independent longitudinal and cross-cultural validation.

## Supporting information

Multimodal_MRI-20260909.docx

## Data Availability

All data produced in the present study are available upon reasonable request to the authors

## Ethics approval and consent to participate

The Ethics Committee of the University of Electronic Science and Technology of China approved this study (approval number: 30850). All procedures involving human participants were conducted in accordance with the Declaration of Helsinki. We obtained written informed consent from all participants before they participated in the study.

## Conflict of interest

The authors declare no competing interests.

## Funding

This study was funded by the Basic Research Projects for Central Universities, Medical-Engineering Cross-Disciplinary Projects grant No. ZYGX2025YGLH010 (by Dr. Yuting Wang)

## Author contributions

**Conceptualization:** Hongzhou Wu and Michael Maes. **Methodology:** Hongzhou Wu, Bharat B. Biswal, Benjamin Klugah-Brown, and Michael Maes. **Formal analysis:** Hongzhou Wu, Chenghui Yang, Ying He, Xianfeng Qu, Yingqiang Zhang, Jinming Xiao, and Chengxiao Yang. **Investigation:** Hongzhou Wu, Chenghui Yang, Ying He, Xianfeng Qu, Elijah Agoalikum, and Yingqiang Zhang. **Data curation:** Hongzhou Wu, Chenghui Yang, Ying He, Elijah Agoalikum, and Yingqiang Zhang. **Visualization:** Hongzhou Wu, Abbas F. Almulla. **Writing – original draft:** Hongzhou Wu, Elijah Agoalikum, and Michael Maes. **Writing – review & editing:** Hongzhou Wu, Chenghui Yang, Ying He, Xianfeng Qu, Abbas F. Almulla, Yingqiang Zhang, Jinming Xiao, Chengxiao Yang, Drozdstoj Stoyanov, Andre F Carvalho, Bharat B. Biswal, Stefania Ferraro, Benjamin Klugah-Brown, Licia Pacheco Luna, Yuting Wang, Elijah Agoalikum, and Michael Maes. **Supervision:** Bharat B. Biswal, Benjamin Klugah-Brown, and Michael Maes. **Project administration:** Elijah Agoalikum and Michael Maes.

All authors contributed to the critical revision of the manuscript for important intellectual content, approved the final version of the manuscript, and agreed to be accountable for their contributions to the work.

## Acknowledgements

The authors sincerely thank all participants for their time and contribution to this study. We also thank the clinical, laboratory, and MRI staff of the Sichuan Provincial Center for Mental Health, Sichuan Provincial People’s Hospital, for their assistance with participant recruitment, clinical assessments, biospecimen collection and processing, and MRI data acquisition.

## Data availability

The de-identified data supporting the findings of this study are available from the corresponding author upon reasonable request, subject to applicable ethical, institutional, and data-protection requirements.

## Code availability

The analysis scripts used to generate the principal results reported in this study are available from the corresponding author upon reasonable request.

## Consent for publication

Not applicable. No identifiable individual participant information is presented in this manuscript.

## Notes

### Competing Interest Statement

The authors have declared no competing interest.

### Author Declarations

Ethics Committee of University of Electronic Science and Technology of China gave ethical approval for this work (approval number: 30850).

## Reference

1. Zachar P, Stoyanov D St., Aragona M, Jablensky A, editors. Alternative Perspectives on Psychiatric Validation: DSM, ICD, RDoC, and Beyond. Oxford: Oxford University Press; 2015.

2. Di Nicola V, Stoyanov D. Psychiatry in Crisis: At the Crossroads of Social Sciences, the Humanities, and Neuroscience. Cham: Springer; 2021.

3. Stoyanov D, Maes MH. How to construct neuroscience-informed psychiatric classification? Towards nomothetic networks psychiatry. World J Psychiatry. 2021;11:1–12.

4. Maes M, Vojdani A, Galecki P, Kanchanatawan B. How to Construct a Bottom-Up Nomothetic Network Model and Disclose Novel Nosological Classes by Integrating Risk Resilience and Adverse Outcome Pathways with the Phenome of Schizophrenia. Brain Sci. 2020;10:645.

5. Maes M, Moraes JB, Bonifacio KL, Barbosa DS, Vargas HO, Michelin AP, et al. Towards a new model and classification of mood disorders based on risk resilience, neuro-affective toxicity, staging, and phenome features using the nomothetic network psychiatry approach. Metab Brain Dis. 2021;36:509–521.

6. Xiang J, Ran J, Zhu P, Wang Y, Deng X, Yang B, et al. Transdiagnostic mapping of common and specific regional homogeneity alterations across affective and psychotic disorders. Neuropsychopharmacology. 2026. June 2026. 10.1038/s41386-026-02469-0.

7. Huang CC, Luo Q, Palaniyappan L, Yang AC, Hung CC, Chou KH, et al. Transdiagnostic and Illness-Specific Functional Dysconnectivity Across Schizophrenia, Bipolar Disorder, and Major Depressive Disorder. Biol Psychiatry Cogn Neurosci Neuroimaging. 2020;5:542–553.

8. Gong J, Wang J, Qiu S, Chen P, Luo Z, Wang J, et al. Common and distinct patterns of intrinsic brain activity alterations in major depression and bipolar disorder: voxel-based meta-analysis. Transl Psychiatry. 2020;10.

9. Kühn S, Gallinat J. Resting-state brain activity in schizophrenia and major depression: A quantitative meta-analysis. Schizophr Bull. 2013;39:358–365.

10. Insel T, Cuthbert B, Garvey M, Heinssen R, Pine DS, Quinn K, et al. Research Domain Criteria (RDoC): Toward a new classification framework for research on mental disorders. American Journal of Psychiatry. 2010;167:748–751.

11. Parkes L, Satterthwaite TD, Bassett DS. Towards precise resting-state fMRI biomarkers in psychiatry: synthesizing developments in transdiagnostic research, dimensional models of psychopathology, and normative neurodevelopment. Curr Opin Neurobiol. 2020;65:120–128.

12. Zou QH, Zhu CZ, Yang Y, Zuo XN, Long XY, Cao QJ, et al. An improved approach to detection of amplitude of low-frequency fluctuation (ALFF) for resting-state fMRI: Fractional ALFF. J Neurosci Methods. 2008;172:137–141.

13. Zang Y, Jiang T, Lu Y, He Y, Tian L. Regional homogeneity approach to fMRI data analysis. Neuroimage. 2004;22:394–400.

14. Biswal B, Zerrin Yetkin F, Haughton VM, Hyde JS. Functional connectivity in the motor cortex of resting human brain using echo planar mri. Magn Reson Med. 1995;34:537–541.

15. Zuo XN, Ehmke R, Mennes M, Imperati D, Castellanos FX, Sporns O, et al. Network centrality in the human functional connectome. Cerebral Cortex. 2012;22:1862–1875.

16. Maes M, Almulla AF, You Z, Zhang Y. Neuroimmune, metabolic and oxidative stress pathways in major depressive disorder. Nat Rev Neurol. 2025;21:473–489.

17. Maes M, Vojdani A, Sirivichayakul S, Barbosa DS, Kanchanatawan B. Inflammatory and Oxidative Pathways Are New Drug Targets in Multiple Episode Schizophrenia and Leaky Gut, Klebsiella pneumoniae, and C1q Immune Complexes Are Additional Drug Targets in First Episode Schizophrenia. Mol Neurobiol. 2021;58:3319–3334.

18. Maes M, Sirivichayakul S, Matsumoto AK, Michelin AP, de Oliveira Semeão L, de Lima Pedrão JV, et al. Lowered Antioxidant Defenses and Increased Oxidative Toxicity Are Hallmarks of Deficit Schizophrenia: a Nomothetic Network Psychiatry Approach. Mol Neurobiol. 2020;57:4578–4597.

19. Morris G, Puri BK, Bortolasci CC, Carvalho A, Berk M, Walder K, et al. The role of high-density lipoprotein cholesterol, apolipoprotein A and paraoxonase-1 in the pathophysiology of neuroprogressive disorders. Neurosci Biobehav Rev. 2021;125:244–263.

20. Nunes SOV, Piccoli De Melo LG, Pizzo De Castro MR, Barbosa DS, Vargas HO, Berk M, et al. Atherogenic index of plasma and atherogenic coefficient are increased in major depression and bipolar disorder, especially when comorbid with tobacco use disorder. J Affect Disord. 2015;172:55–62.

21. Onen S, Taymur I. Evidence for the atherogenic index of plasma as a potential biomarker for cardiovascular disease in schizophrenia. Journal of Psychopharmacology. 2021;35:1120–1126.

22. Tien YT, Wang LJ, Lee Y, Lin PY, Hung CF, Chong MY, et al. Comparative predictive efficacy of atherogenic indices on metabolic syndrome in patients with schizophrenia. Schizophr Res. 2023;262:95–101.

23. Maes M, Zhou B, Jirakran K, Vasupanrajit A, Boonchaya-Anant P, Tunvirachaisakul C, et al. Towards a major methodological shift in depression research by assessing continuous scores of recurrence of illness, lifetime and current suicidal behaviors and phenome features. J Affect Disord. 2024;350:728–740.

24. Castro-Gomez S, Heneka MT. Innate immune activation in neurodegenerative diseases. Immunity. 2024;57:790–814.

25. Miller AH, Raison CL. The role of inflammation in depression: From evolutionary imperative to modern treatment target. Nat Rev Immunol. 2016;16:22–34.

26. Wu H, Xiao J, Agoalikum E, Baig TI, Becker B, Ferraro S, et al. Multi-Indicator Entropy Hub Score: A quantitative approach to hub analysis in brain networks. Neuroimage. 2026;329.

27. Wu H, Yang Z, Cao Q, Wang P, Biswal BB, Klugah-Brown B. MQGA: A quantitative analysis of brain network hubs using multi-graph theoretical indices. Neuroimage. 2024;303.

28. Tomasi D, Wang GJ, Volkow ND. Energetic cost of brain functional connectivity. Proc Natl Acad Sci U S A. 2013;110:13642–13647.

29. Crossley NA, Mechelli A, Scott J, Carletti F, Fox PT, Mcguire P, et al. The hubs of the human connectome are generally implicated in the anatomy of brain disorders. Brain. 2014;137:2382–2395.

30. Yu T, Zhao G, Sun Y, Lu Z, Liao Y, Yuan R, et al. The multimodal neuroimaging signatures and gene expression profiles for adverse childhood experiences. BMC Med. 2025;23.

31. Watters ER, Aloe AM, Wojciak AS. Examining the Associations Between Childhood Trauma, Resilience, and Depression: A Multivariate Meta-Analysis. Trauma Violence Abuse. 2023;24:231–244.

32. Chen SS, He Y, Xie GD, Chen LR, Zhang TT, Yuan MY, et al. Relationships among adverse childhood experience patterns, psychological resilience, self-esteem and depressive symptoms in Chinese adolescents: A serial multiple mediation model. Prev Med (Baltim). 2022;154.

33. Yan CG, Wang ZH, Han LKM, Alexander N, Alnæs D, Başgöze Z, et al. Vertex-wise cortical abnormalities in major depressive disorder from 64 cohorts from the DIRECT and ENIGMA MDD consortia. Nature Mental Health. 2026. July 2026. 10.1038/s44220-026-00667-9.

34. Georgiadis F, Larivière S, Glahn D, Hong LE, Kochunov P, Mowry B, et al. Connectome architecture shapes large-scale cortical alterations in schizophrenia: a worldwide ENIGMA study. Mol Psychiatry. 2024;29:1869–1881.

35. Magioncalda P, Yadav A, Martino M. An umbrella review of neuroimaging studies and conceptual framework linking pathophysiology and psychopathology in schizophrenia. Nature Mental Health. 2025;3:1241–1255.

36. American Psychiatric Association. Diagnostic and Statistical Manual of Mental Disorders. 5th ed. Arlington, VA: American Psychiatric Association; 2013.

37. Harry Stack S. The Psychiatric Interview. 1970.

38. Sheehan D V., Lecrubier Y, Sheehan KH, Amorim P, Janavs J, Weiller E, et al. The Mini-International Neuropsychiatric Interview (M.I.N.I.): The development and validation of a structured diagnostic psychiatric interview for DSM-IV and ICD-10. Journal of Clinical Psychiatry, vol. 59, 1998. p. 22–33.

39. Hamilton M. A rating scale for depression. J Neurol Neurosurg Psychiatry. 1960;23:56–62.

40. Kay SR, Fiszbein A, Opler LA. The positive and negative syndrome scale (PANSS) for schizophrenia. Schizophr Bull. 1987;13:261–276.

41. Beck AT, Steer RA, Brown G. Beck Depression Inventory–II. PsycTESTS Dataset. 2011.

42. Spielberger C, Gorsuch R, Lushene R. Manual for the State-Trait Anxiety Inventory. Https://WwwSemanticscholarOrg/Paper/Manual-for-the-State-Trait-Anxiety-Inventory-Spielberger-Gorsuch/E6d09d04fc8737094c193da471e2a50a809f77d4. 1970.

43. Bernstein DP, Stein JA, Newcomb MD, Walker E, Pogge D, Ahluvalia T, et al. Development and validation of a brief screening version of the Childhood Trauma Questionnaire. Child Abuse Negl. 2003;27:169–190.

44. Connor KM, Davidson JRT. Development of a new Resilience scale: The Connor-Davidson Resilience scale (CD-RISC). Depress Anxiety. 2003;18:76–82.

45. Yan CG, Wang X Di, Zuo XN, Zang YF. DPABI: Data Processing & Analysis for (Resting-State) Brain Imaging. Neuroinformatics. 2016;14:339–351.

46. Wang J, Wang X, Xia M, Liao X, Evans A, He Y. GRETNA: a graph theoretical network analysis toolbox for imaging connectomics. Front Hum Neurosci. 2015;9.

47. Wager TD, Davidson ML, Hughes BL, Lindquist MA, Ochsner KN. Prefrontal-Subcortical Pathways Mediating Successful Emotion Regulation. Neuron. 2008;59:1037–1050.

48. Wager TD, Waugh CE, Lindquist M, Noll DC, Fredrickson BL, Taylor SF. Brain mediators of cardiovascular responses to social threat. Part I: Reciprocal dorsal and ventral sub-regions of the medial prefrontal cortex and heart-rate reactivity. Neuroimage. 2009;47:821–835.

49. Troyanskaya O, Cantor M, Sherlock G, Brown P, Hastie T, Tibshirani R, et al. Missing value estimation methods for DNA microarrays. Bioinformatics. 2001;17:520–525.

50. Galindo-Prieto B, Eriksson L, Trygg J. Variable influence on projection (VIP) for orthogonal projections to latent structures (OPLS). J Chemom. 2014;28:623–632.

51. Thévenot EA, Roux A, Xu Y, Ezan E, Junot C. Analysis of the Human Adult Urinary Metabolome Variations with Age, Body Mass Index, and Gender by Implementing a Comprehensive Workflow for Univariate and OPLS Statistical Analyses. J Proteome Res. 2015;14:3322–3335.

52. Breiman L. Random forests. Mach Learn. 2001;45:5–32.

53. Lewis MJ, Spiliopoulou A, Goldmann K, Pitzalis C, McKeigue P, Barnes MR. nestedcv: an R package for fast implementation of nested cross-validation with embedded feature selection designed for transcriptomics and high-dimensional data. Bioinformatics Advances. 2023;3:vbad048.

54. Kuhn M. Building predictive models in R using the caret package. J Stat Softw. 2008;28:1–26.

55. Hair JF, Howard MC, Nitzl C. Assessing measurement model quality in PLS-SEM using confirmatory composite analysis. J Bus Res. 2020;109:101–110.

56. Henseler J, Ringle CM, Sarstedt M. A new criterion for assessing discriminant validity in variance-based structural equation modeling. J Acad Mark Sci. 2015;43:115–135.

57. Goodkind M, Eickhoff SB, Oathes DJ, Jiang Y, Chang A, Jones-Hagata LB, et al. Identification of a common neurobiological substrate for mental Illness. JAMA Psychiatry. 2015;72:305–315.

58. McTeague LM, Goodkind MS, Etkin A. Transdiagnostic impairment of cognitive control in mental illness. J Psychiatr Res. 2016;83:37–46.

59. Craig AD. How do you feel - now? The anterior insula and human awareness. Nat Rev Neurosci. 2009;10:59–70.

60. Damasio A, Carvalho GB. The nature of feelings: Evolutionary and neurobiological origins. Nat Rev Neurosci. 2013;14:143–152.

61. Ferraro S, Klugah-Brown B, Tench CR, Bazinet V, Bore MC, Nigri A, et al. The central autonomic system revisited – Convergent evidence for a regulatory role of the insular and midcingulate cortex from neuroimaging meta-analyses. Neurosci Biobehav Rev. 2022;142.

62. Ford JM, Dierks T, Fisher DJ, Herrmann CS, Hubl D, Kindler J, et al. Neurophysiological studies of auditory verbal hallucinations. Schizophr Bull. 2012;38:715–723.

63. Sliz D, Hayley S. Major depressive disorder and alterations in insular cortical activity: a review of current functional magnetic imaging research. Front Hum Neurosci. 2012;6:323.

64. Wu N, Liu T, Tian M, Liu C, Ma S, Cao H, et al. Albumin, an interesting and functionally diverse protein, varies from ‘native’ to ‘effective’ (Review). Mol Med Rep. 2024;29.

65. Maes M, Almulla AF, Stoyanov D, Zhang Y. Unifying the hallmarks of major depression through neuroimmune-metabolic-oxidative (NIMETOX) dysregulation: a mechanistic systems framework. Cell Mol Immunol. 2026;23:933–957.

66. Li S, Hu N, Zhang W, Tao B, Dai J, Gong Y, et al. Dysconnectivity of Multiple Brain Networks in Schizophrenia: A Meta-Analysis of Resting-State Functional Connectivity. Front Psychiatry. 2019;10:482.

67. Guo L, Ma J, Cai M, Zhang M, Xu Q, Wang H, et al. Transcriptional signatures of the whole-brain voxel-wise resting-state functional network centrality alterations in schizophrenia. Schizophrenia (Heidelberg, Germany). 2023;9:87.

68. Cao J, Chen X, Chen J, Ai M, Gan Y, He J, et al. The Association Between Resting State Functional Connectivity and the Trait of Impulsivity and Suicidal Ideation in Young Depressed Patients With Suicide Attempts. Front Psychiatry. 2021;12.

69. Bryant RA, Breukelaar IA, Williamson T, Felmingham K, Williams LM, Korgaonkar MS. The neural connectome of suicidality in adults with mood and anxiety disorders. Nature Mental Health. 2024;2:1342–1349.

70. Geffen T, Hardikar S, Smallwood J, Kaliuzhna M, Carruzzo F, Böge K, et al. Striatal Functional Hypoconnectivity in Patients with Schizophrenia Suffering from Negative Symptoms, Longitudinal Findings. Schizophr Bull. 2024;50:1337–1348.

