## Supplementary material for "A Systems Biology Model Linking Peripheral NIMETOX Biomarkers to Multilevel Brain Structural and Functional Alterations Across Major Depression and Schizophrenia": Multimodal_MRI-20260909.docx

**ELECTRONIC SUPPLEMENTARY FILE (ESF)**

**ESF, Table 1.** Methods used to assay the biomarkers in the present study.

|  | **Assay** | **Method and kit** | **Equipment** |
| --- | --- | --- | --- |
| 1 | Albumin | Immunoturbidimetric assay (DIAYS DIAGNOSTIC SYSTEM (SHANGHAI) CO., LTD). Sensitivity 0.03 g/L, intra-assay and inter-assay analytical coefficients of variation (CVs) of 1.96% and 0.67%, respectively. | Fully automated biochemical analyzer (ADVIA 2400, Siemens Healthcare Diagnostic Inc) |
| 2 | Triglycerides (TG) | GPO-PAP method (Gcell, Beijing Strong Biotechnologies, Inc.), with the intra-assay and inter-assay analytical CVs of 3.0% and 7.0%, respectively. | Fully automated biochemical analyzer (ADVIA 2400, Siemens Healthcare Diagnostic Inc) |
| 3 | Total cholesterol (TC) | Cholesterol oxidase-peroxidase-aminoantipyrine-phenol (CHOD-PAP) method (Beijing Strong Biotechnologies, Inc.) with the intra-assay and inter-assay CVs of < 4% and < 6%, respectively. | Fully automated biochemical analyzer (ADVIA 2400, Siemens Healthcare Diagnostic Inc) |
| 4 | Low-density lipoprotein cholesterol (LDL-C) | Direct method-surfactant clearance method (Beijing Strong Biotechnologies, Inc.) with the intra-assay and inter-assay CVs of < 3% and < 10%, respectively. | Fully automated biochemical analyzer (ADVIA 2400, Siemens Healthcare Diagnostic Inc) |
| 5 | High-density lipoprotein cholesterol (HDL-C) | Direct method-select inhibition method (Beijing Strong Biotechnologies, Inc.) with the intra-assay and inter-assay CVs of < 4% and < 10%, respectively. | Fully automated biochemical analyzer (ADVIA 2400, Siemens Healthcare Diagnostic Inc) |
| 6 | LDL-C / HDL-C | Castelli risk index 2 | See above |
| 7 | Apolipoprotein A1 (ApoA1) | Immunoturbidimetric method (Beijing Strong Biotechnologies, Inc.) with the intra-assay and inter-assay CVs of < 3% and < 10%, respectively. | Fully automated biochemical analyzer (ADVIA 2400, Siemens Healthcare Diagnostic Inc) |
| 8 | Free cholesterol (FC) | CHOD-PAP method (mlbio, China) with the intra-assay and inter-assay CVs of < 3% and < 5%, respectively. | Fully automated biochemical analyzer (ADVIA 2400, Siemens Healthcare Diagnostic Inc) |

**ESF, Table 2**. Sociodemographic and clinical characteristics of patients with major depressive disorder (MDD), schizophrenia (SCZ), and healthy controls (HC).

| Variables | HCᵃ (n = 26) | MDDᵇ (n = 38) | SCZᶜ (n = 29) | Test statistic (F/KW/χ²) | df | *p-value* | Multiple comparisons |
| --- | --- | --- | --- | --- | --- | --- | --- |
| Age (years) | 33.9 ± 14.0 | 28.3 ± 9.5 | 33.0 ± 12.6 | 2.16 | 2/90 | 0.122 | — |
| Female/male ratio | 16/10 | 25/13 | 17/12 | 0.37 | 2 | 0.831 | __ |
| Education (years) | 12.8 ± 3.5 | 14.2 ± 2.2 | 13.0 ± 3.1 | 1.85 | 2/84 | 0.164 | — |
| BMI (kg/m²) | 25.1 ± 4.9 | 22.7 ± 3.8 | 21.8 ± 3.4 | 4.93 | 2/86 | 0.009 | a > b,c |
| Sexual abuse score | 5.3 ± 0.7 | 7.1 ± 3.2 | 7.2 ± 3.2 | 2.89 | 2/81 | 0.061 | — |
| ACEs score | 31.0 ± 7.5 | 43.5 ± 13.2 | 37.0 ± 10.9 | 9.32 | 2/80 | <0.001 | b > c > a |
| PC-RISK | 0.74 ± 0.60 | -0.82 ± 0.56 | 0.21 ± 0.93 | 40.65 | 2/90 | <0.001 | a > c > b |
| BDI-II | 4.31 ± 4.17 | 36.81 ± 12.62 | 12.04 ± 9.64 | - | - | <0.001 | b > c > a |
| State STAI | 32.38 ± 9.94 | 57.80 ± 13.56 | 40.42 ± 15.01 | - | - | <0.001 | b > c > a |
| OSOM (z score) | -0.96 ± 0.27 | 1.09 ± 0.53 | -0.23 ± 0.61 | - | - | <0.001 | b > c > a |
| Positive symptoms (pos) | 0.00 ± 0.00 | 12.21 ± 7.06 | 24.69 ± 9.40 | - | - | <0.001 | c > b > a |
| Negative symptoms (neg) | 0.00 ± 0.00 | 13.34 ± 8.13 | 23.72 ± 10.57 | - | - | <0.001 | c > b > a |
| OSOP (z score) | -1.10 ± 0.00 | 0.00 ± 0.617 | 0.99 ± 0.71 | - | - | <0.001 | c > b > a |
| Current suicidal ideation (z score) | -0.88 ± 0.08 | -0.01 ± 0.91 | 0.82 ± 0.79 | - | - | <0.001 | c > b > a |
| Total suicidal behaviors (z score) | -1.12 ± 0.18 | 0.29 ± 0.86 | 0.67 ± 0.71 | - | - | <0.001 | c > b > a |

Values are mean ± SD. F: analysis of variance (ANOVA), KW: Kruskal-Wallis non-parametric ANOVA; χ²: analysis of contingency tables. Group codes: a = HC, b = MDD, and c = SCZ. Multiple comparisons are based on LSD tests. Only p < 0.05 pairwise differences are shown when the omnibus ANOVA is significant; > indicates the group with the higher mean.

**ESF, Table 3.** Plasma biomarker characteristics of patients with major depressive disorder (MDD), schizophrenia (SCZ) and healthy controls (HC)

| Variables | HCᵃ (n = 26) | MDDᵇ (n = 38) | SCZᶜ (n = 29) | | Test statistic (F/U/χ²) | df | *p-value* | Multiple comparisons |
| --- | --- | --- | --- | --- | --- | --- | --- | --- |
| Albumin (g/L) | 45.33 ± 2.18 | 42.33 ± 3.20 | 43.13 ± 3.71 | 7.023 | | 2/86 | 0.001 | a > b,c |
| Triglycerides (mmol/L) | 1.18 ± 0.67 | 1.42 ± 0.76 | 1.36 ± 0.63 | 0.983 | | 2/86 | 0.378 | — |
| Total cholesterol (mmol/L) | 4.80 ± 0.98 | 4.87 ± 0.80 | 4.42 ± 0.92 | 2.120 | | 2/86 | 0.126 | — |
| Free cholesterol (mmol/L) | 0.385 (0.097) | 0.406 (0.127) | 0.417 (0.143) | 0.426 | | 2/86 | 0.655 | --- |
| HDL-C (mmol/L) | 1.37 ± 0.27 | 1.14 ± 0.33 | 1.22 ± 0.34 | 3.990 | | 2/86 | 0.022 | a > b |
| LDL-C (mmol/L) | 2.72 ± 0.89 | 2.98 ± 0.71 | 2.54 ± 0.75 | 2.508 | | 2/86 | 0.087 | — |
| VLDL-C (mmol/L) | 0.70 ± 0.30 | 0.75 ± 0.20 | 0.66 ± 0.22 | 1.230 | | 2/86 | 0.297 | — |
| LDL-C/HDL-C ratio | 2.10 ± 0.93 | 2.87 ± 1.16 | 2.22 ± 0.86 | 5.26 | | 2/86 | 0.007 | b > a,c |
| ApoA1 (g/L) | 1.73 ± 0.27 | 1.378 ± 0.30 | 1.51 ± 0.30 | 11.10 | | 2/86 | <0.001 | a > b,c |
| NAPR (z score) | 0.73 ± 0.72 | -0.49 ± 0.84 | -0.08 ± 1.03 | 14.675 | | 2/86 | <0.001 | a > b,c |
| Atherogenic index of plasma (z score) | -0.38 ± 0.85 | 0.25 ± 1.09 | 0.05 ± 0.93 | 3.192 | | 2/86 | 0.046 | b > a |

Shown are mean values ± SD. GLM analysis showed that age (F=2.55, df=10/73, p=0.011) and sex (F=2.93, df=10/73, p=0.004) has significant effects, whereas body mass index (F=1.75, df=10/73, p=0.086) and smoking (F=1.52, df=10/73, p=0.149) had not. Covarying did not change any of the above contrasts. The adjusted z values can be requested from the authors. Header n denotes the maximum available group size; the valid n varies across measures because of missing data. Group codes: a = HC, b = MDD, and c = SCZ. Multiple comparisons are based on LSD tests. Only p < 0.05 pairwise differences are shown when the omnibus ANOVA is significant; > indicates the group with the higher mean. Note: TG: triglycerides; TC: total cholesterol; HDL-C: high-density lipoprotein cholesterol; LDL-C: low-density lipoprotein cholesterol; VLDL-C: very low-density lipoprotein cholesterol; ApoA1: apolipoprotein A1; NAPR: z unit composite of z albumin + z ApoA1).

**ESF, Table 4**. Mediation analysis of GMV, fALFF, and clinical measurements.

| **X** | **M** | **Y** | **N** | **a** | **p_a** | **b** | **p_b** | **cPrime** | **p_cPrime** | **c** | **p_c** | **ab** | **p_ab** |
| --- | --- | --- | --- | --- | --- | --- | --- | --- | --- | --- | --- | --- | --- |
| **GMV02** | **fALFF02** | **PANSS_pos** | **87** | **1.696186** | **0.017205** | **-0.67341** | **0.000815** | **0.42** | **0.800562** | **-0.74441** | **0.556985** | **-1.16441** | **0.005999** |
| **GMV02** | **fALFF02** | **OSOI** | **87** | **1.690876** | **0.018368** | **-0.91044** | **8.37E-05** | **-1.22332** | **0.310511** | **-2.7671** | **0.029938** | **-1.54378** | **0.007213** |
| **GMV02** | **fALFF02** | **OSOP** | **87** | **1.68268** | **0.018214** | **-0.67653** | **0.000966** | **-0.36251** | **0.827561** | **-1.52092** | **0.244091** | **-1.15842** | **0.007744** |
| **GMV02** | **fALFF02** | **PC_RISC** | **87** | **1.687032** | **0.01513** | **0.480178** | **0.010398** | **2.155816** | **0.082854** | **2.973932** | **0.021279** | **0.818116** | **0.011274** |
| **GMV02** | **fALFF02** | **PANSS_neg** | **87** | **1.6832** | **0.020208** | **-0.60715** | **0.003517** | **-1.09021** | **0.453738** | **-2.12283** | **0.083671** | **-1.03262** | **0.013819** |
| **GMV02** | **fALFF02** | **PC_QoL** | **74** | **1.970357** | **0.023504** | **0.582891** | **0.008378** | **0.690964** | **0.650447** | **1.847201** | **0.192554** | **1.156238** | **0.015347** |
| **GMV02** | **fALFF02** | **CD_RISC** | **81** | **1.821875** | **0.017548** | **12.29292** | **0.014099** | **54.72265** | **0.072286** | **77.38494** | **0.016319** | **22.66229** | **0.015758** |
| **GMV02** | **fALFF02** | **OSOM** | **87** | **1.692383** | **0.018761** | **-0.75418** | **0.000103** | **-1.16453** | **0.315784** | **-2.4374** | **0.050784** | **-1.27287** | **0.016147** |
| **GMV02** | **fALFF02** | **total_SB** | **87** | **1.696335** | **0.020449** | **-0.69118** | **0.007554** | **1.166931** | **0.401034** | **-0.01384** | **0.944278** | **-1.18077** | **0.018095** |
| **GMV02** | **fALFF02** | **PC_Impuse** | **87** | **1.683936** | **0.017859** | **-0.59015** | **0.003022** | **-2.98081** | **0.016002** | **-3.98153** | **0.000487** | **-1.00072** | **0.019415** |
| **GMV02** | **fALFF02** | **BSI2_NoPlan** | **80** | **1.815954** | **0.021746** | **-3.16757** | **0.006617** | **-17.142** | **0.051803** | **-22.9555** | **0.004649** | **-5.81356** | **0.023638** |
| **GMV02** | **fALFF02** | **Current_SI** | **87** | **1.684798** | **0.018428** | **-0.57352** | **0.005655** | **1.655346** | **0.239358** | **0.672507** | **0.676146** | **-0.98284** | **0.024597** |
| **GMV02** | **fALFF02** | **BSI2_MI** | **79** | **1.867576** | **0.018594** | **-2.14331** | **0.006508** | **-11.6212** | **0.013287** | **-15.6079** | **0.000315** | **-3.98679** | **0.025195** |
| **GMV02** | **fALFF02** | **SA** | **82** | **1.822643** | **0.016531** | **-10.0209** | **0.009659** | **-26.167** | **0.190817** | **-44.0213** | **0.04649** | **-17.8544** | **0.047482** |
| **GMV03** | **fALFF03** | **OSOI** | **87** | **2.273576** | **0.000803** | **-0.91253** | **1.44E-05** | **-1.90004** | **0.121748** | **-3.94956** | **0.000921** | **-2.04952** | **0.000771** |
| **GMV03** | **fALFF03** | **OSOM** | **87** | **2.277146** | **0.000319** | **-0.81763** | **0.000273** | **-1.01964** | **0.546425** | **-2.89523** | **0.04198** | **-1.87559** | **0.001266** |
| **GMV03** | **fALFF03** | **TA** | **81** | **2.171849** | **0.00094** | **-9.30453** | **0.005067** | **-5.08189** | **0.868384** | **-26.0038** | **0.153174** | **-20.9219** | **0.007202** |
| **GMV03** | **fALFF03** | **PANSS_pos** | **87** | **2.26695** | **0.000291** | **-0.698** | **0.007542** | **-0.61852** | **0.671764** | **-2.16138** | **0.127267** | **-1.54286** | **0.007982** |
| **GMV03** | **fALFF03** | **SA** | **82** | **2.187805** | **0.000626** | **-10.9431** | **0.010441** | **-16.8532** | **0.505892** | **-41.1465** | **0.07021** | **-24.2933** | **0.018019** |
| **GMV03** | **fALFF03** | **total_SB** | **87** | **2.282654** | **0.000351** | **-0.52697** | **0.015918** | **-0.97269** | **0.50934** | **-2.17668** | **0.099185** | **-1.20399** | **0.018434** |
| **GMV03** | **fALFF03** | **OSOP** | **87** | **2.274349** | **0.000289** | **-0.64833** | **0.018458** | **-1.49464** | **0.27564** | **-2.9274** | **0.025895** | **-1.43276** | **0.021679** |
| **GMV03** | **fALFF03** | **BSI2_AI** | **80** | **1.963142** | **0.003559** | **-1.21758** | **0.012706** | **-6.31591** | **0.148905** | **-8.70493** | **0.036381** | **-2.38901** | **0.024736** |
| **GMV03** | **fALFF03** | **CTQ_SF_EA** | **83** | **2.178615** | **0.00068** | **-2.44353** | **0.020738** | **-8.66985** | **0.136549** | **-14.0533** | **0.006106** | **-5.38344** | **0.026055** |
| **GMV03** | **fALFF03** | **CTQ_SF_Total** | **81** | **2.304531** | **0.000426** | **-7.22631** | **0.016532** | **-46.189** | **0.01157** | **-62.8242** | **0.000384** | **-16.6353** | **0.033925** |
| **GMV03** | **fALFF03** | **Current_SI** | **87** | **2.274432** | **0.000324** | **-0.47479** | **0.037986** | **1.214359** | **0.443194** | **0.154366** | **0.930324** | **-1.05999** | **0.049746** |
| **GMV04** | **fALFF04** | **total_SB** | **87** | **1.629027** | **0.001639** | **-0.92584** | **2.15E-05** | **2.778672** | **0.016498** | **1.271779** | **0.230357** | **-1.50689** | **0.001278** |
| **GMV04** | **fALFF04** | **OSOI** | **87** | **1.624749** | **0.002455** | **-0.78422** | **0.000238** | **-1.28911** | **0.217685** | **-2.56406** | **0.006744** | **-1.27495** | **0.002139** |
| **GMV04** | **fALFF04** | **PANSS_pos** | **87** | **1.634037** | **0.00215** | **-0.78085** | **0.00097** | **0.738633** | **0.482888** | **-0.52705** | **0.541142** | **-1.26568** | **0.003801** |
| **GMV04** | **fALFF04** | **PANSS_neg** | **87** | **1.626608** | **0.002793** | **-0.68662** | **0.00104** | **0.293094** | **0.775135** | **-0.82439** | **0.403564** | **-1.11749** | **0.003894** |
| **GMV04** | **fALFF04** | **Current_SI** | **87** | **1.614617** | **0.003825** | **-0.84978** | **2.37E-05** | **3.538286** | **0.003329** | **2.165046** | **0.059241** | **-1.37324** | **0.0039** |
| **GMV04** | **fALFF04** | **OSOP** | **87** | **1.625861** | **0.002985** | **-0.7743** | **0.000383** | **0.544788** | **0.602156** | **-0.71113** | **0.443487** | **-1.25592** | **0.003939** |
| **GMV04** | **fALFF04** | **CTQ_SF_Total** | **81** | **1.639011** | **0.004793** | **-6.97491** | **0.013866** | **-13.127** | **0.397025** | **-24.8667** | **0.125833** | **-11.7397** | **0.022308** |
| **GMV04** | **fALFF04** | **OSOM** | **87** | **1.623189** | **0.003373** | **-0.52211** | **0.016419** | **-2.00446** | **0.104389** | **-2.85845** | **0.011193** | **-0.85399** | **0.027962** |
| **GMV04** | **fALFF04** | **CTQ_SF_PN** | **83** | **1.593675** | **0.008679** | **-1.67472** | **0.01917** | **5.038164** | **0.160218** | **2.313916** | **0.444262** | **-2.72425** | **0.03559** |
| **GMV07** | **fALFF07** | **OSOM** | **87** | **2.276928** | **0.000887** | **-0.86535** | **0.000203** | **-1.57058** | **0.123778** | **-3.51691** | **0.002267** | **-1.94633** | **0.000168** |
| **GMV07** | **fALFF07** | **PANSS_pos** | **87** | **2.279217** | **0.002083** | **-0.65692** | **0.000396** | **0.930883** | **0.567426** | **-0.60661** | **0.571775** | **-1.53749** | **0.000301** |
| **GMV07** | **fALFF07** | **OSOI** | **87** | **2.282961** | **0.004076** | **-0.95237** | **0.000154** | **-1.1433** | **0.267085** | **-3.31922** | **0.005818** | **-2.17592** | **0.00053** |
| **GMV07** | **fALFF07** | **OSOP** | **87** | **2.270435** | **0.002364** | **-0.62891** | **0.001258** | **0.101841** | **0.972998** | **-1.35356** | **0.264971** | **-1.4554** | **0.001085** |
| **GMV07** | **fALFF07** | **SA** | **82** | **2.376419** | **0.001481** | **-11.6763** | **0.00113** | **-28.8219** | **0.134928** | **-56.289** | **0.005611** | **-27.4671** | **0.003663** |
| **GMV07** | **fALFF07** | **total_SB** | **87** | **2.284897** | **0.001615** | **-0.52986** | **0.003466** | **0.513721** | **0.702442** | **-0.69649** | **0.553996** | **-1.21021** | **0.005349** |
| **GMV07** | **fALFF07** | **TA** | **81** | **2.355404** | **0.001494** | **-8.45619** | **0.002108** | **-27.278** | **0.054583** | **-47.0195** | **0.002633** | **-19.7415** | **0.005468** |
| **GMV07** | **fALFF07** | **PANSS_neg** | **87** | **2.27295** | **0.001525** | **-0.54106** | **0.005935** | **-0.6722** | **0.639253** | **-1.92345** | **0.151909** | **-1.25125** | **0.006777** |
| **GMV07** | **fALFF07** | **CTQ_SF_Total** | **81** | **2.391395** | **0.000754** | **-7.85847** | **0.007483** | **-7.69872** | **0.72339** | **-26.2373** | **0.141693** | **-18.5385** | **0.011795** |
| **GMV07** | **fALFF07** | **BDI2** | **81** | **2.440095** | **0.000816** | **-10.2644** | **0.011079** | **-35.5257** | **0.056046** | **-59.98** | **0.004642** | **-24.4544** | **0.020699** |
| **GMV07** | **fALFF07** | **CTQ_SF_EN** | **82** | **2.361848** | **0.000783** | **-2.91787** | **0.014855** | **1.543042** | **0.852626** | **-5.30052** | **0.4323** | **-6.84356** | **0.020963** |
| **GMV07** | **fALFF07** | **PC_Impuse** | **87** | **2.263501** | **0.0008** | **-0.47419** | **0.02077** | **-3.30203** | **0.004533** | **-4.36635** | **0.000784** | **-1.06432** | **0.029337** |
| **GMV07** | **fALFF07** | **Neuroticism** | **87** | **2.272437** | **0.0008** | **-0.5034** | **0.016699** | **-2.6372** | **0.012699** | **-3.7473** | **0.000568** | **-1.11009** | **0.034094** |
| **GMV07** | **fALFF07** | **CTQ_SF_EA** | **83** | **2.400237** | **0.000758** | **-2.17702** | **0.023639** | **-2.90622** | **0.574213** | **-8.00096** | **0.159289** | **-5.09474** | **0.037543** |
| **GMV07** | **fALFF07** | **Brooding** | **87** | **2.277443** | **0.000981** | **-0.4585** | **0.020202** | **-1.59357** | **0.184418** | **-2.62286** | **0.034322** | **-1.02929** | **0.039633** |
| **GMV07** | **fALFF07** | **RRS** | **82** | **2.435716** | **0.000858** | **-8.41741** | **0.029324** | **-36.9976** | **0.062956** | **-57.3386** | **0.002729** | **-20.341** | **0.045145** |
| **GMV09** | **fALFF09** | **OSOI** | **87** | **0.954426** | **0.004801** | **-0.83241** | **0.001832** | **-1.41003** | **0.145169** | **-2.19318** | **0.015203** | **-0.78315** | **0.018339** |
| **GMV09** | **fALFF09** | **PC_RISC** | **87** | **0.95045** | **0.004434** | **0.566944** | **0.049031** | **1.773828** | **0.071922** | **2.311643** | **0.014346** | **0.537815** | **0.030946** |
| **GMV09** | **fALFF09** | **OSOM** | **87** | **0.959903** | **0.004881** | **-0.68812** | **0.007625** | **-1.28701** | **0.140238** | **-1.93823** | **0.024868** | **-0.65121** | **0.031389** |
| **GMV09** | **fALFF09** | **OSOP** | **87** | **0.9576** | **0.006032** | **-0.68203** | **0.00986** | **-0.70027** | **0.496802** | **-1.34619** | **0.145283** | **-0.64592** | **0.038393** |
| **GMV09** | **fALFF09** | **PC_Impuse** | **87** | **0.953425** | **0.004332** | **-0.72102** | **0.014316** | **-2.41446** | **0.007294** | **-3.09221** | **0.000492** | **-0.67775** | **0.038536** |
| **GMV09** | **fALFF09** | **TA** | **81** | **1.006694** | **0.002558** | **-8.88489** | **0.017617** | **-9.68713** | **0.380669** | **-18.5069** | **0.091881** | **-8.81982** | **0.041568** |
| **GMV09** | **fALFF09** | **PANSS_neg** | **87** | **0.95389** | **0.005104** | **-0.67928** | **0.013702** | **-1.07731** | **0.273696** | **-1.71271** | **0.06133** | **-0.63539** | **0.04693** |
| **GMV10** | **fALFF10** | **CTQ_SF_EA** | **83** | **1.701846** | **0.013988** | **-2.79323** | **0.006676** | **-4.25672** | **0.455548** | **-9.03599** | **0.089234** | **-4.77928** | **0.008705** |
| **GMV10** | **fALFF10** | **OSOP** | **87** | **1.534612** | **0.015281** | **-0.71542** | **0.001041** | **-0.81365** | **0.542991** | **-1.91252** | **0.132776** | **-1.09887** | **0.013136** |
| **GMV10** | **fALFF10** | **PANSS_neg** | **87** | **1.534004** | **0.017505** | **-0.7541** | **0.00098** | **-0.75027** | **0.622605** | **-1.90013** | **0.164712** | **-1.14986** | **0.013432** |
| **GMV10** | **fALFF10** | **OSOI** | **87** | **1.533095** | **0.015132** | **-0.80057** | **0.000414** | **-2.55856** | **0.028312** | **-3.7859** | **0.00244** | **-1.22734** | **0.01408** |
| **GMV10** | **fALFF10** | **CTQ_SF_PA** | **83** | **1.68423** | **0.013972** | **-1.94203** | **0.009402** | **-0.5634** | **0.820947** | **-3.80857** | **0.254609** | **-3.24516** | **0.017273** |
| **GMV10** | **fALFF10** | **PC_QoL** | **74** | **1.516911** | **0.029743** | **0.54715** | **0.023806** | **2.867659** | **0.058574** | **3.662927** | **0.018762** | **0.795268** | **0.018597** |
| **GMV10** | **fALFF10** | **PANSS_pos** | **87** | **1.537878** | **0.019017** | **-0.60625** | **0.005556** | **-0.79214** | **0.554956** | **-1.73873** | **0.155514** | **-0.94659** | **0.021072** |
| **GMV10** | **fALFF10** | **CTQ_SF_Total** | **81** | **1.519748** | **0.029953** | **-9.83901** | **0.000843** | **-4.0318** | **0.781973** | **-18.9185** | **0.256484** | **-14.8867** | **0.022993** |
| **GMV10** | **fALFF10** | **CD_RISC** | **81** | **1.540256** | **0.027628** | **11.24204** | **0.04181** | **99.91205** | **0.008188** | **116.1999** | **0.003694** | **16.28787** | **0.023469** |
| **GMV10** | **fALFF10** | **PC_RISC** | **87** | **1.541169** | **0.014966** | **0.456088** | **0.046543** | **3.454032** | **0.023025** | **4.123986** | **0.01159** | **0.669954** | **0.024629** |
| **GMV10** | **fALFF10** | **SA** | **82** | **1.664187** | **0.014038** | **-11.0109** | **0.006616** | **-40.8996** | **0.028193** | **-59.136** | **0.002739** | **-18.2365** | **0.024836** |
| **GMV10** | **fALFF10** | **Neuroticism** | **87** | **1.538606** | **0.017679** | **-0.63967** | **0.004226** | **-3.49021** | **0.001241** | **-4.46486** | **0.000135** | **-0.97465** | **0.027608** |
| **GMV10** | **fALFF10** | **PC_Impuse** | **87** | **1.542837** | **0.018107** | **-0.62328** | **0.013613** | **-3.60447** | **0.001479** | **-4.55693** | **8.35E-05** | **-0.95245** | **0.042183** |
| **GMV10** | **fALFF10** | **TA** | **81** | **1.71086** | **0.013861** | **-7.18507** | **0.022112** | **-32.1956** | **0.045251** | **-44.6081** | **0.005855** | **-12.4125** | **0.046376** |
| **GMV10** | **fALFF10** | **OSOM** | **87** | **1.5418** | **0.015287** | **-0.5836** | **0.016554** | **-3.03429** | **0.014328** | **-3.92841** | **0.001488** | **-0.89411** | **0.046408** |
| **GMV11** | **fALFF11** | **Current_SI** | **87** | **1.837027** | **0.007235** | **-0.79139** | **5.83E-05** | **1.951503** | **0.172175** | **0.487156** | **0.761856** | **-1.46435** | **0.003289** |
| **GMV11** | **fALFF11** | **total_SB** | **87** | **1.840364** | **0.006626** | **-0.9944** | **0.000288** | **0.84796** | **0.515233** | **-0.97231** | **0.493824** | **-1.82027** | **0.003461** |
| **GMV11** | **fALFF11** | **PANSS_pos** | **87** | **1.828994** | **0.006106** | **-0.80799** | **6.18E-05** | **1.355539** | **0.215743** | **-0.13925** | **0.921966** | **-1.49479** | **0.003615** |
| **GMV11** | **fALFF11** | **OSOP** | **87** | **1.836875** | **0.005167** | **-0.75857** | **0.000105** | **0.410108** | **0.701813** | **-0.99517** | **0.407518** | **-1.40528** | **0.003837** |
| **GMV11** | **fALFF11** | **OSOI** | **87** | **1.839813** | **0.00713** | **-0.874** | **0.000142** | **-1.22405** | **0.235206** | **-2.82881** | **0.02104** | **-1.60476** | **0.007384** |
| **GMV11** | **fALFF11** | **PANSS_neg** | **87** | **1.837164** | **0.007027** | **-0.6338** | **0.002352** | **-0.49813** | **0.756498** | **-1.66938** | **0.220891** | **-1.17126** | **0.008832** |
| **GMV11** | **fALFF11** | **OSOM** | **87** | **1.839527** | **0.005406** | **-0.64337** | **0.005911** | **-2.43454** | **0.027646** | **-3.6043** | **0.001936** | **-1.16976** | **0.015723** |
| **GMV11** | **fALFF11** | **CTQ_SF_Total** | **81** | **1.776343** | **0.016596** | **-8.0375** | **0.003134** | **-7.04748** | **0.630443** | **-21.044** | **0.194496** | **-13.9965** | **0.027287** |
| **GMV11** | **fALFF11** | **SA** | **82** | **1.700484** | **0.022669** | **-10.8195** | **0.005247** | **-6.48885** | **0.64311** | **-24.7089** | **0.190818** | **-18.2201** | **0.031349** |
| **GMV11** | **fALFF11** | **CTQ_SF_PN** | **83** | **1.706312** | **0.017884** | **-1.50819** | **0.010062** | **-3.27961** | **0.411755** | **-5.82584** | **0.159359** | **-2.54623** | **0.042575** |
| **GMV11** | **fALFF11** | **TA** | **81** | **1.749418** | **0.016302** | **-6.94847** | **0.01882** | **-3.81805** | **0.74986** | **-15.8238** | **0.309092** | **-12.0058** | **0.045976** |

Model: X -> M -> Y mediation with covariates controlled in all regressions; a: X -> M; b: M -> Y controlling for X; cPrime: Direct effect X -> Y controlling for M; c: Total effect X -> Y without M; ab: Indirect/mediation effect = a*b; Estimate: stats.mean from MediationToolbox bootstrap results; matches pathway diagram values; SE: stats.ste, i.e., SD of bootstrap coefficient distribution; p: Bias-corrected bootstrap p value returned by MediationToolbox; Covariates: Sex, Age, BMI; Bootstrap samples: 10000;

**ESF, Table 5.** Mediation analysis of GMV, ReHo, and clinical measurements.

| **X** | **M** | **Y** | **N** | **a** | **p_a** | **b** | **p_b** | **cPrime** | **p_cPrime** | **c** | **p_c** | **ab** | **p_ab** |
| --- | --- | --- | --- | --- | --- | --- | --- | --- | --- | --- | --- | --- | --- |
| **GMV03** | **ReHo03** | **OSOI** | **87** | **1.755215** | **0.008432** | **-0.919882** | **8.28E-05** | **-2.354676** | **0.057115** | **-3.984898** | **0.001057** | **-1.630222** | **0.003424** |
| **GMV03** | **ReHo03** | **PANSS_pos** | **87** | **1.755298** | **0.009204** | **-0.730632** | **0.001099** | **-0.871574** | **0.563756** | **-2.150067** | **0.134657** | **-1.278493** | **0.008736** |
| **GMV03** | **ReHo03** | **OSOM** | **87** | **1.752752** | **0.009024** | **-0.690832** | **0.005368** | **-1.65945** | **0.307283** | **-2.899431** | **0.043733** | **-1.239981** | **0.011878** |
| **GMV03** | **ReHo03** | **OSOP** | **87** | **1.750593** | **0.013977** | **-0.759739** | **0.000263** | **-1.591084** | **0.220533** | **-2.917894** | **0.024963** | **-1.326809** | **0.012577** |
| **GMV03** | **ReHo03** | **PANSS_neg** | **87** | **1.740515** | **0.012526** | **-0.710375** | **0.002424** | **-2.110574** | **0.11084** | **-3.344944** | **0.013187** | **-1.23437** | **0.017797** |
| **GMV03** | **ReHo03** | **total_SB** | **87** | **1.755464** | **0.010804** | **-0.591225** | **0.004596** | **-1.152561** | **0.362314** | **-2.179867** | **0.104607** | **-1.027306** | **0.018519** |
| **GMV03** | **ReHo03** | **SA** | **82** | **1.857301** | **0.012668** | **-10.7939** | **0.015405** | **-20.35349** | **0.407121** | **-41.11464** | **0.071475** | **-20.76115** | **0.026945** |
| **GMV03** | **ReHo03** | **TA** | **81** | **1.863015** | **0.010849** | **-7.83835** | **0.029739** | **-10.3379** | **0.623444** | **-25.64394** | **0.182886** | **-15.30603** | **0.049955** |
| **GMV04** | **ReHo04** | **PANSS_pos** | **87** | **1.525675** | **0.006373** | **-0.747811** | **0.000259** | **0.627006** | **0.567871** | **-0.517939** | **0.567505** | **-1.144945** | **0.0043** |
| **GMV04** | **ReHo04** | **OSOI** | **87** | **1.530187** | **0.006082** | **-0.657726** | **0.002095** | **-1.520318** | **0.168818** | **-2.544063** | **0.011542** | **-1.023745** | **0.004508** |
| **GMV04** | **ReHo04** | **OSOP** | **87** | **1.514636** | **0.00538** | **-0.711676** | **0.001327** | **0.371009** | **0.709813** | **-0.715974** | **0.467435** | **-1.086984** | **0.005085** |
| **GMV04** | **ReHo04** | **Current_SI** | **87** | **1.516213** | **0.008319** | **-0.621039** | **0.004309** | **3.086714** | **0.008915** | **2.15103** | **0.053635** | **-0.935684** | **0.016334** |
| **GMV04** | **ReHo04** | **total_SB** | **87** | **1.522576** | **0.007164** | **-0.642082** | **0.007095** | **2.248955** | **0.057053** | **1.277069** | **0.245351** | **-0.971887** | **0.017772** |
| **GMV04** | **ReHo04** | **PANSS_neg** | **87** | **1.519708** | **0.007432** | **-0.605992** | **0.009211** | **0.124151** | **0.930146** | **-0.812676** | **0.422015** | **-0.936827** | **0.018838** |
| **GMV04** | **ReHo04** | **PANSS_all** | **61** | **1.778125** | **0.008337** | **-19.70106** | **0.01826** | **45.45939** | **0.333571** | **9.911461** | **0.798667** | **-35.54793** | **0.018927** |
| **GMV07** | **ReHo07** | **OSOI** | **87** | **1.848024** | **0.000828** | **-1.053681** | **3.16E-05** | **-1.334725** | **0.249148** | **-3.292401** | **0.005682** | **-1.957676** | **0.000228** |
| **GMV07** | **ReHo07** | **OSOP** | **87** | **1.860295** | **0.00091** | **-0.911539** | **0.000221** | **0.346445** | **0.818684** | **-1.358075** | **0.275851** | **-1.70452** | **0.00037** |
| **GMV07** | **ReHo07** | **PANSS_pos** | **87** | **1.846192** | **0.000757** | **-0.835582** | **0.000114** | **0.933935** | **0.46734** | **-0.611389** | **0.571574** | **-1.545324** | **0.00056** |
| **GMV07** | **ReHo07** | **PANSS_neg** | **87** | **1.858371** | **0.00201** | **-0.893859** | **0.000205** | **-0.277677** | **0.850658** | **-1.953731** | **0.134841** | **-1.676053** | **0.000929** |
| **GMV07** | **ReHo07** | **OSOM** | **87** | **1.842798** | **0.001119** | **-0.748118** | **0.000341** | **-2.117041** | **0.080656** | **-3.506514** | **0.002656** | **-1.389473** | **0.001013** |
| **GMV07** | **ReHo07** | **total_SB** | **87** | **1.849906** | **0.001418** | **-0.702611** | **0.001263** | **0.634994** | **0.683428** | **-0.683246** | **0.575796** | **-1.318241** | **0.001268** |
| **GMV07** | **ReHo07** | **Current_SI** | **87** | **1.84576** | **0.000765** | **-0.544589** | **0.01234** | **2.446037** | **0.123735** | **1.410414** | **0.258941** | **-1.035623** | **0.014824** |
| **GMV07** | **ReHo07** | **CTQ_SF_Total** | **81** | **1.736306** | **0.002546** | **-7.547642** | **0.019368** | **-13.27358** | **0.482292** | **-26.09933** | **0.148705** | **-12.82575** | **0.038666** |
| **GMV07** | **ReHo07** | **TA** | **81** | **1.547304** | **0.00518** | **-7.234227** | **0.025147** | **-35.83861** | **0.023967** | **-47.25743** | **0.002222** | **-11.41883** | **0.048037** |
| **GMV08** | **ReHo08** | **CD_RISC** | **81** | **1.414363** | **0.029603** | **14.78068** | **0.002503** | **25.48163** | **0.414317** | **46.18154** | **0.133902** | **20.69991** | **0.016852** |
| **GMV08** | **ReHo08** | **PC_RISC** | **87** | **1.245943** | **0.039578** | **0.662305** | **0.00058** | **0.894607** | **0.432118** | **1.717898** | **0.153403** | **0.823291** | **0.026657** |
| **GMV08** | **ReHo08** | **OSOP** | **87** | **1.259366** | **0.041416** | **-0.810748** | **2.28E-05** | **-0.606227** | **0.619622** | **-1.631797** | **0.159047** | **-1.02557** | **0.046005** |
| **GMV09** | **ReHo09** | **BDI2** | **81** | **1.167568** | **0.003465** | **-17.44934** | **6.96E-05** | **-19.53412** | **0.221869** | **-39.78231** | **0.006957** | **-20.24818** | **0.003753** |
| **GMV09** | **ReHo09** | **TA** | **81** | **1.141775** | **0.005189** | **-14.63244** | **0.000101** | **-1.774241** | **0.905402** | **-18.42588** | **0.101456** | **-16.65164** | **0.004622** |
| **GMV09** | **ReHo09** | **PC_RISC** | **87** | **1.120821** | **0.004633** | **0.850469** | **0.001589** | **1.345536** | **0.110772** | **2.297396** | **0.015424** | **0.95186** | **0.004913** |
| **GMV09** | **ReHo09** | **SA** | **82** | **1.124673** | **0.002884** | **-14.54363** | **0.000405** | **-16.91099** | **0.253034** | **-33.12765** | **0.01336** | **-16.21667** | **0.006407** |
| **GMV09** | **ReHo09** | **CD_RISC** | **81** | **1.14894** | **0.004815** | **19.72902** | **0.005394** | **31.24514** | **0.156042** | **53.87872** | **0.026707** | **22.63358** | **0.006785** |
| **GMV09** | **ReHo09** | **OSOM** | **87** | **1.116112** | **0.004556** | **-1.158413** | **4.25E-05** | **-0.61717** | **0.48355** | **-1.909726** | **0.030125** | **-1.292555** | **0.007542** |
| **GMV09** | **ReHo09** | **OSOI** | **87** | **1.118467** | **0.005817** | **-1.173218** | **3.91E-05** | **-0.861409** | **0.373242** | **-2.182447** | **0.018796** | **-1.321039** | **0.008245** |
| **GMV09** | **ReHo09** | **RRS** | **82** | **1.104642** | **0.00591** | **-15.68328** | **0.000213** | **-6.720375** | **0.662734** | **-23.89143** | **0.108884** | **-17.17106** | **0.008611** |
| **GMV09** | **ReHo09** | **total_SB** | **87** | **1.124812** | **0.004509** | **-0.787914** | **0.000103** | **-0.792832** | **0.455444** | **-1.673514** | **0.083973** | **-0.880683** | **0.008966** |
| **GMV09** | **ReHo09** | **PC_Impuse** | **87** | **1.124912** | **0.004012** | **-0.961558** | **0.002199** | **-2.043297** | **0.019585** | **-3.117795** | **0.000227** | **-1.074498** | **0.01088** |
| **GMV09** | **ReHo09** | **Neuroticism** | **87** | **1.120263** | **0.004048** | **-0.866051** | **0.001532** | **-0.952266** | **0.272325** | **-1.911007** | **0.018574** | **-0.958741** | **0.011711** |
| **GMV09** | **ReHo09** | **BSI2_MI** | **79** | **1.105152** | **0.007758** | **-3.450324** | **0.000628** | **-9.069619** | **0.002985** | **-12.86737** | **8.28E-05** | **-3.797748** | **0.01348** |
| **GMV09** | **ReHo09** | **OSOP** | **87** | **1.129073** | **0.003746** | **-0.756891** | **0.003382** | **-0.4569** | **0.717313** | **-1.327885** | **0.155998** | **-0.870985** | **0.014709** |
| **GMV09** | **ReHo09** | **BSI2_NoPlan** | **80** | **1.110502** | **0.006913** | **-5.38621** | **0.004788** | **-9.829901** | **0.110605** | **-15.83812** | **0.01123** | **-6.008222** | **0.016017** |
| **GMV09** | **ReHo09** | **PC_QoL** | **74** | **0.96279** | **0.020682** | **0.808507** | **0.003787** | **0.57304** | **0.529208** | **1.336305** | **0.140939** | **0.763264** | **0.016393** |
| **GMV09** | **ReHo09** | **BSI2_total** | **79** | **1.107455** | **0.008005** | **-10.15813** | **0.004902** | **-24.3184** | **0.021638** | **-35.58367** | **0.00053** | **-11.26527** | **0.016797** |
| **GMV09** | **ReHo09** | **Brooding** | **87** | **1.123025** | **0.006042** | **-0.628619** | **0.00575** | **-0.492504** | **0.634718** | **-1.198052** | **0.15365** | **-0.705548** | **0.017016** |
| **GMV09** | **ReHo09** | **PANSS_pos** | **87** | **1.115995** | **0.004277** | **-0.750234** | **0.007151** | **0.025289** | **0.976395** | **-0.82977** | **0.371669** | **-0.855059** | **0.023065** |
| **GMV09** | **ReHo09** | **CTQ_SF_SA** | **82** | **1.094967** | **0.005648** | **-2.241196** | **0.00537** | **-3.553543** | **0.198007** | **-5.979962** | **0.026021** | **-2.426419** | **0.024373** |
| **GMV09** | **ReHo09** | **PANSS_neg** | **87** | **1.122505** | **0.003301** | **-0.674861** | **0.010351** | **-0.939408** | **0.370829** | **-1.708077** | **0.060438** | **-0.768669** | **0.033035** |
| **GMV10** | **ReHo10** | **CD_RISC** | **81** | **2.082871** | **0.00338** | **16.20572** | **0.000289** | **82.93745** | **0.022299** | **116.525** | **0.005774** | **33.5876** | **0.002464** |
| **GMV10** | **ReHo10** | **SA** | **82** | **2.088934** | **0.003412** | **-12.82688** | **2.79E-05** | **-32.38112** | **0.10308** | **-59.31479** | **0.003115** | **-26.93367** | **0.003278** |
| **GMV10** | **ReHo10** | **TA** | **81** | **2.18031** | **0.002164** | **-9.075592** | **0.000992** | **-24.26816** | **0.132834** | **-44.37569** | **0.006911** | **-20.10753** | **0.005127** |
| **GMV10** | **ReHo10** | **OSOI** | **87** | **1.799886** | **0.008328** | **-1.021291** | **0.000116** | **-1.93171** | **0.117892** | **-3.779032** | **0.003027** | **-1.847322** | **0.005707** |
| **GMV10** | **ReHo10** | **PANSS_neg** | **87** | **1.803732** | **0.007611** | **-0.851435** | **0.000283** | **-0.417055** | **0.769684** | **-1.931446** | **0.151551** | **-1.514391** | **0.005994** |
| **GMV10** | **ReHo10** | **OSOP** | **87** | **1.793507** | **0.007491** | **-0.833782** | **0.000475** | **-0.430124** | **0.754697** | **-1.915527** | **0.122129** | **-1.485402** | **0.006242** |
| **GMV10** | **ReHo10** | **BDI2** | **81** | **2.214054** | **0.002013** | **-10.78809** | **0.004212** | **-40.98921** | **0.06872** | **-65.2471** | **0.001489** | **-24.25789** | **0.007137** |
| **GMV10** | **ReHo10** | **CTQ_SF_EA** | **83** | **2.0981** | **0.004073** | **-2.406048** | **0.00509** | **-3.788856** | **0.559889** | **-8.901605** | **0.110786** | **-5.112748** | **0.007337** |
| **GMV10** | **ReHo10** | **OSOM** | **87** | **1.792471** | **0.007916** | **-0.789813** | **9.71E-05** | **-2.483936** | **0.055716** | **-3.92104** | **0.000576** | **-1.437104** | **0.007438** |
| **GMV10** | **ReHo10** | **PANSS_pos** | **87** | **1.787323** | **0.007992** | **-0.731625** | **0.000491** | **-0.437348** | **0.74444** | **-1.743268** | **0.147692** | **-1.30592** | **0.007512** |
| **GMV10** | **ReHo10** | **PC_QoL** | **74** | **2.01002** | **0.006228** | **0.508173** | **0.007396** | **2.658344** | **0.081268** | **3.65708** | **0.008994** | **0.998736** | **0.00971** |
| **GMV10** | **ReHo10** | **CTQ_SF_PA** | **83** | **2.079998** | **0.003123** | **-1.509418** | **0.008775** | **-0.616933** | **0.811933** | **-3.766721** | **0.280965** | **-3.149788** | **0.009976** |
| **GMV10** | **ReHo10** | **PC_RISC** | **87** | **1.79672** | **0.013694** | **0.709319** | **0.000268** | **2.805793** | **0.041163** | **4.094547** | **0.008406** | **1.288755** | **0.010284** |
| **GMV10** | **ReHo10** | **Neuroticism** | **87** | **1.790173** | **0.009556** | **-0.609581** | **0.001277** | **-3.373105** | **0.003999** | **-4.465361** | **8.08E-05** | **-1.092256** | **0.012071** |
| **GMV10** | **ReHo10** | **CTQ_SF_Total** | **81** | **2.024813** | **0.003651** | **-7.356378** | **0.00476** | **-4.386692** | **0.796396** | **-19.02127** | **0.249825** | **-14.63458** | **0.013775** |
| **GMV10** | **ReHo10** | **PC_Impuse** | **87** | **1.798675** | **0.010928** | **-0.568495** | **0.003669** | **-3.527415** | **0.005548** | **-4.566026** | **8.21E-05** | **-1.03861** | **0.014803** |
| **GMV10** | **ReHo10** | **BSI2_MI** | **79** | **2.058632** | **0.004411** | **-2.079638** | **0.011288** | **-14.28358** | **0.00252** | **-18.65169** | **5.03E-05** | **-4.36811** | **0.019105** |
| **GMV10** | **ReHo10** | **total_SB** | **87** | **1.804302** | **0.011267** | **-0.477978** | **0.011815** | **-1.133679** | **0.358286** | **-2.011443** | **0.084151** | **-0.877765** | **0.026045** |
| **GMV10** | **ReHo10** | **RRS** | **82** | **2.061954** | **0.003533** | **-8.884882** | **0.018128** | **-50.23518** | **0.022236** | **-69.16883** | **0.000525** | **-18.93365** | **0.030228** |
| **GMV10** | **ReHo10** | **BSI2_total** | **79** | **2.052848** | **0.002182** | **-5.657809** | **0.02643** | **-47.38408** | **0.002495** | **-59.2365** | **4.9E-05** | **-11.85242** | **0.035169** |
| **GMV11** | **ReHo11** | **total_SB** | **87** | **1.578767** | **0.024372** | **-0.844876** | **0.000232** | **0.364777** | **0.793837** | **-0.974582** | **0.501679** | **-1.339359** | **0.012339** |
| **GMV11** | **ReHo11** | **Current_SI** | **87** | **1.563232** | **0.025038** | **-0.697444** | **0.000272** | **1.591207** | **0.260772** | **0.476068** | **0.753393** | **-1.115138** | **0.014389** |
| **GMV11** | **ReHo11** | **PANSS_pos** | **87** | **1.5629** | **0.026773** | **-0.777784** | **1E-04** | **1.08891** | **0.364978** | **-0.136285** | **0.922323** | **-1.225195** | **0.015933** |
| **GMV11** | **ReHo11** | **PANSS_neg** | **87** | **1.565903** | **0.02488** | **-0.749833** | **0.000183** | **-0.513542** | **0.705215** | **-1.682328** | **0.228692** | **-1.168786** | **0.018019** |
| **GMV11** | **ReHo11** | **OSOP** | **87** | **1.56806** | **0.031029** | **-0.799776** | **0.000168** | **0.296349** | **0.83397** | **-0.961093** | **0.475514** | **-1.257442** | **0.019692** |
| **GMV11** | **ReHo11** | **OSOI** | **87** | **1.568528** | **0.026986** | **-0.816758** | **6.91E-05** | **-1.553711** | **0.143664** | **-2.829893** | **0.024372** | **-1.276182** | **0.024786** |
| **GMV11** | **ReHo11** | **OSOM** | **87** | **1.574953** | **0.026882** | **-0.515024** | **0.008564** | **-2.814909** | **0.007582** | **-3.615413** | **0.001575** | **-0.800503** | **0.045607** |

Model: X -> M -> Y mediation with covariates controlled in all regressions; a: X -> M; b: M -> Y controlling for X; cPrime: Direct effect X -> Y controlling for M; c: Total effect X -> Y without M; ab: Indirect/mediation effect = a*b; Estimate: stats.mean from MediationToolbox bootstrap results; matches pathway diagram values; SE: stats.ste, i.e., SD of bootstrap coefficient distribution; p: Bias-corrected bootstrap p value returned by MediationToolbox; Covariates: Sex, Age, BMI; Bootstrap samples: 10000;

**ESF, Table 6**. Total indirect effects from PLS-SE

| Indirect paths | **t** | **p** |
| --- | --- | --- |
| **ACEs -> OSOM** | 2.404 | 0.016 |
| **AIP -> Intrinsic functional activity sensorimotor** | -2.073 | 0.038 |
| **AIP -> OSOM** | 2.097 | 0.036 |
| **AIP -> OSOP** | 1.812 | 0.070 |
| **AIP -> PC_RISC** | -2.030 | 0.042 |
| **ANTIOX-INFLAM -> GMV-sensorimotor** | 2.170 | 0.030 |
| **ANTIOX-INFLAM -> Intrinsic functional activity sensorimotor** | 1.929 | 0.054 |
| **ANTIOX-INFLAM -> OSOM** | -2.165 | 0.030 |
| **ANTIOX-INFLAM -> OSOP** | -2.221 | 0.026 |
| **ANTIOX-INFLAM -> PC_RISC** | 1.872 | 0.061 |
| **Age -> Intrinsic functional activity sensorimotor** | -4.364 | <0.001 |
| **Age -> OSOM** | -2.180 | 0.029 |
| **Age -> OSOP** | 3.219 | 0.001 |
| **Age -> PC_RISC** | -3.142 | 0.002 |
| **GMV-sensorimotor -> OSOM** | -3.875 | <0.001 |
| **GMV-sensorimotor -> OSOP** | -3.323 | 0.001 |

**ESF, Table 7.** Specific indirect effects from PLS-SEM

|  | **t** | **p** |
| --- | --- | --- |
| **ANTIOX-INFLAM -> Intrinsic functional activity sensorimotor -> OSOM** | -1.47 | 0.143 |
| **ANTIOX-INFLAM -> Intrinsic functional activity sensorimotor -> OSOP** | -1.96 | 0.050 |
| **ANTIOX-INFLAM -> AIP -> GMV-sensorimotor** | 2.17 | 0.030 |
| **GMV-sensorimotor -> Intrinsic functional activity sensorimotor -> OSOM** | -1.88 | 0.060 |
| **GMV-sensorimotor -> Intrinsic functional activity sensorimotor -> OSOP** | -3.32 | 0.001 |
| **AIP -> GMV-sensorimotor -> Intrinsic functional activity sensorimotor -> OSOP** | 1.81 | 0.070 |
| **ANTIOX-INFLAM -> AIP -> PC_RISC** | 1.50 | 0.134 |
| **AIP -> GMV-sensorimotor -> Intrinsic functional activity sensorimotor -> OSOM** | 1.31 | 0.191 |
| **ANTIOX-INFLAM -> AIP -> GMV-sensorimotor -> PC_RISC** | 1.86 | 0.062 |
| **Age -> GMV-sensorimotor -> Intrinsic functional activity sensorimotor -> OSOP** | 3.22 | 0.001 |
| **Age -> GMV-sensorimotor -> Intrinsic functional activity sensorimotor -> OSOM** | 1.92 | 0.055 |
| **ANTIOX-INFLAM -> AIP -> GMV-sensorimotor -> Intrinsic functional activity sensorimotor** | 1.93 | 0.054 |
| **ANTIOX-INFLAM -> AIP -> GMV-sensorimotor -> PC_RISC -> OSOM** | -1.76 | 0.078 |
| **ACEs -> PC_RISC -> OSOM** | 2.40 | 0.016 |
| **AIP -> PC_RISC -> OSOM** | 1.60 | 0.110 |
| **Age -> PC_RISC -> OSOM** | -3.70 | 0.000 |
| **AIP -> GMV-sensorimotor -> Intrinsic functional activity sensorimotor** | -2.07 | 0.038 |
| **GMV-sensorimotor -> PC_RISC -> OSOM** | -3.36 | 0.001 |
| **Age -> GMV-sensorimotor -> Intrinsic functional activity sensorimotor** | -4.36 | 0.000 |
| **AIP -> GMV-sensorimotor -> PC_RISC** | -2.03 | 0.042 |
| **Age -> GMV-sensorimotor -> PC_RISC** | -3.14 | 0.002 |
| **AIP -> GMV-sensorimotor -> PC_RISC -> OSOM** | 1.88 | 0.061 |
| **Age -> GMV-sensorimotor -> PC_RISC -> OSOM** | 3.05 | 0.002 |
| **ANTIOX-INFLAM -> AIP -> PC_RISC -> OSOM** | -1.46 | 0.144 |
| **ANTIOX-INFLAM -> AIP -> GMV-sensorimotor -> Intrinsic functional activity sensorimotor -> OSOM** | -1.27 | 0.205 |
| **ANTIOX-INFLAM -> AIP -> GMV-sensorimotor -> Intrinsic functional activity sensorimotor -> ZOSOP** | -1.66 | 0.098 |

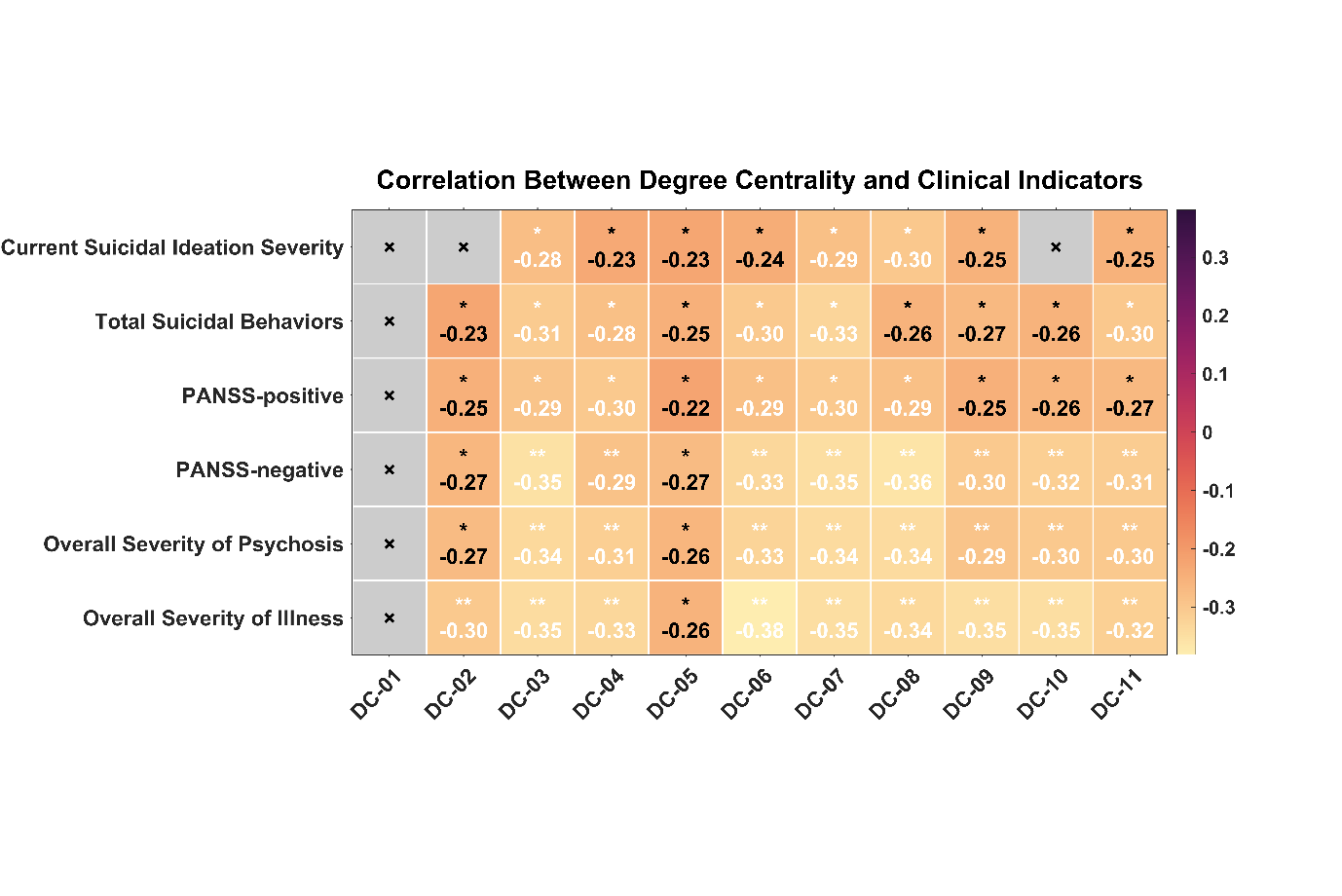

**ESF, Figure 1** Nodal degree centrality associations with dimensional clinical severity. Heatmap of Pearson correlations between degree centrality in ROI01–ROI11 and Current Suicidal Ideation Severity, Total Suicidal Behaviors, PANSS-positive, PANSS-negative, Overall Severity of Psychosis, and Overall Severity of Illness. Cell values are Pearson correlation coefficients; negative values indicate that lower nodal centrality was associated with greater clinical severity. Gray cells marked with “×” denote non-significant associations at the threshold displayed. All significant results passed the FDR correction. * p < 0.05, ** p<0.01, ***p<0.001.

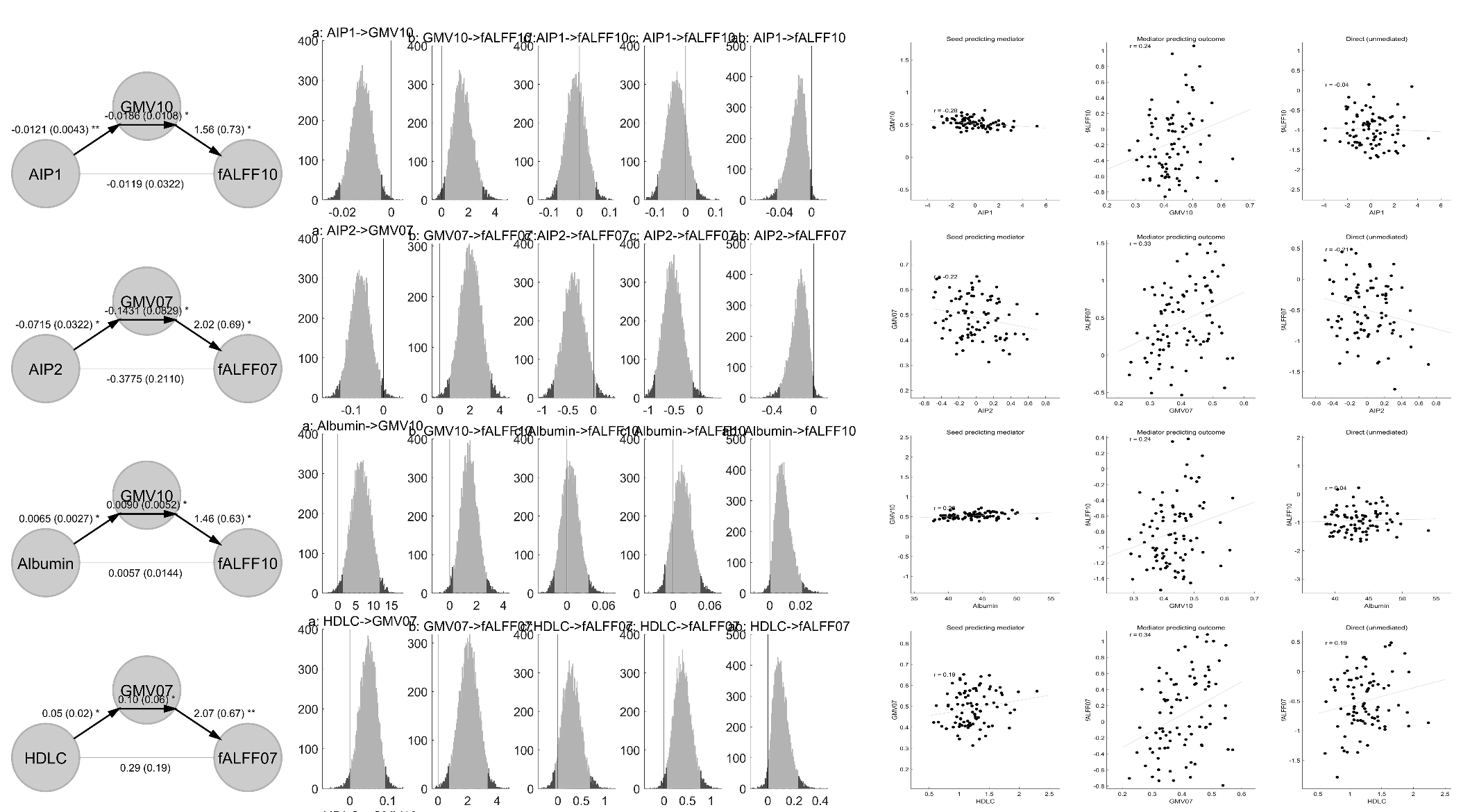

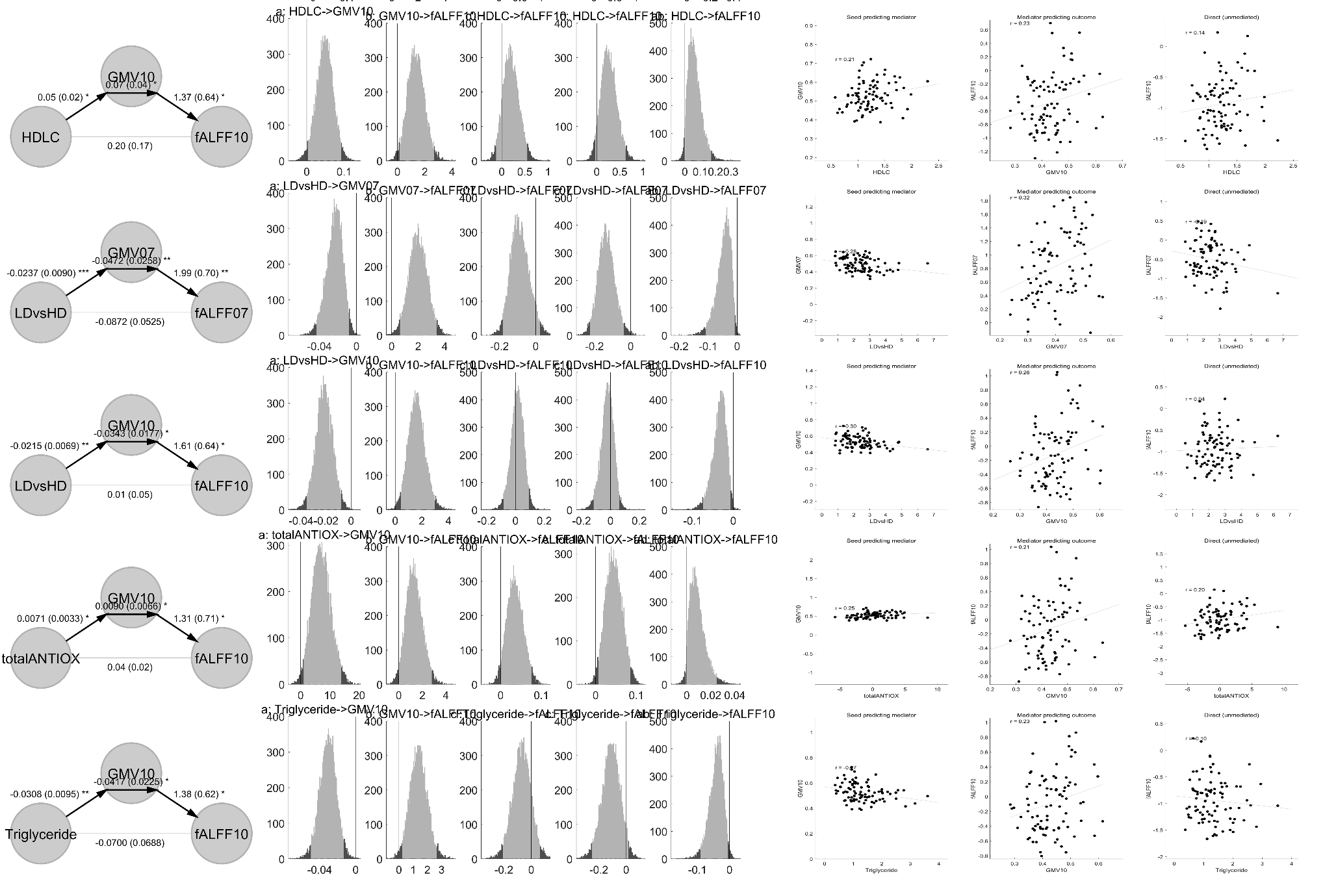

**ESF, Figure 2** Mediation analyses linking peripheral biomarkers to regional gray matter volume (GMV) and fractional amplitude of fALFF. Each row presents one significant mediation model in which a peripheral biomarker was entered as the predictor (X), regional GMV as the mediator (M), and regional fALFF as the outcome (Y). Left panels show the mediation path diagrams, including the X→M (path a), M→Y controlling for X (path b), and direct X→Y effect controlling for M (path c′). Values are regression coefficients with standard errors in parentheses; asterisks indicate statistical significance (P < 0.05, *P < 0.01, **P < 0.001). Middle panels show the bootstrap distributions of the mediation-path coefficients, including the indirect effect (ab). Right panels show the corresponding partial-regression scatterplots for the predictor–mediator association, mediator–outcome association, and direct predictor–outcome association. All mediation models were adjusted for age, sex, and body mass index and were evaluated using 10,000 bootstrap samples. GMV, gray matter volume; fALFF, fractional amplitude of low-frequency fluctuations; HDL-C, high-density lipoprotein cholesterol; LDvsHD, low-density lipoprotein cholesterol/high-density lipoprotein cholesterol ratio; totalANTIOX, NAPR index.

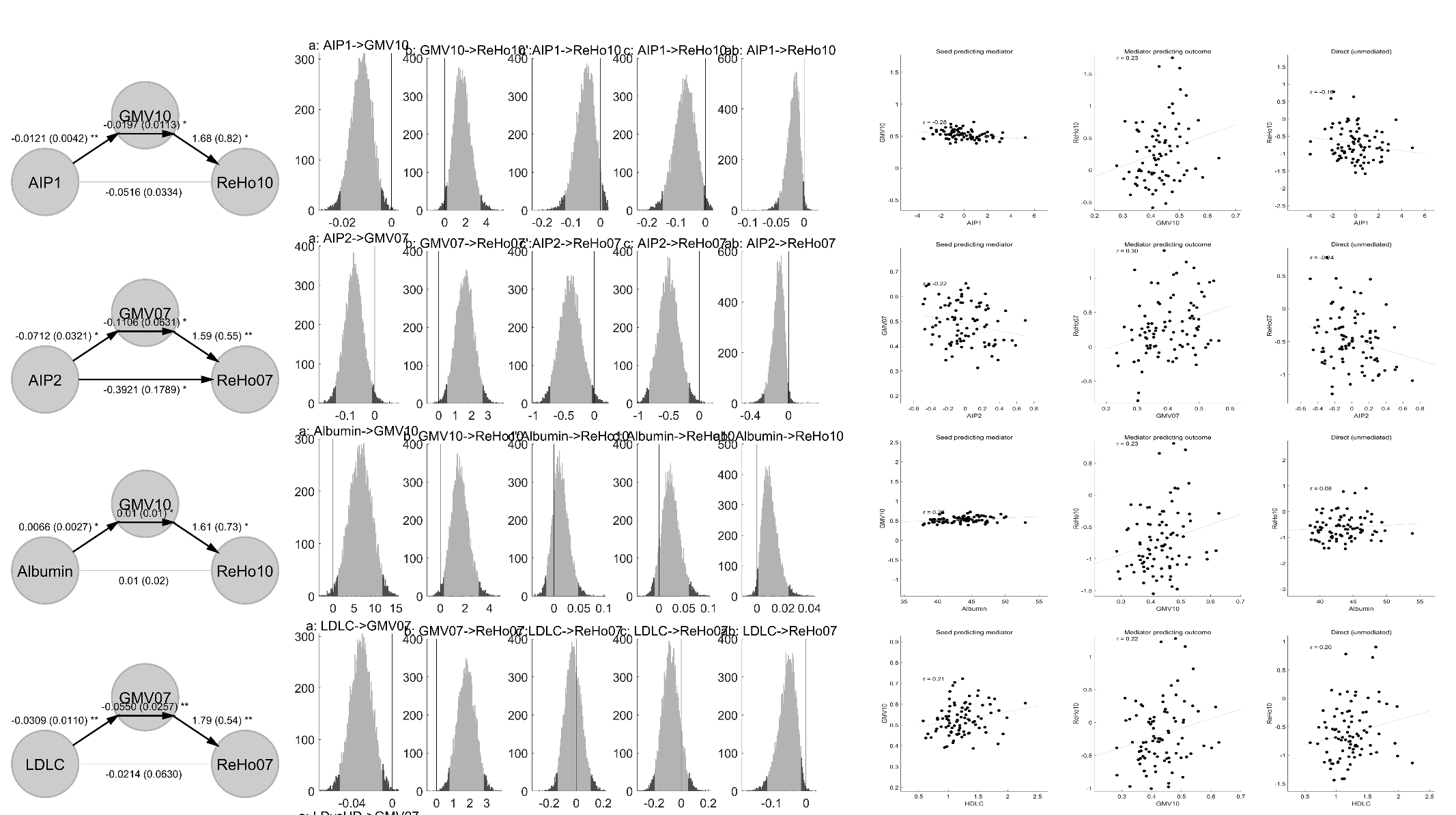

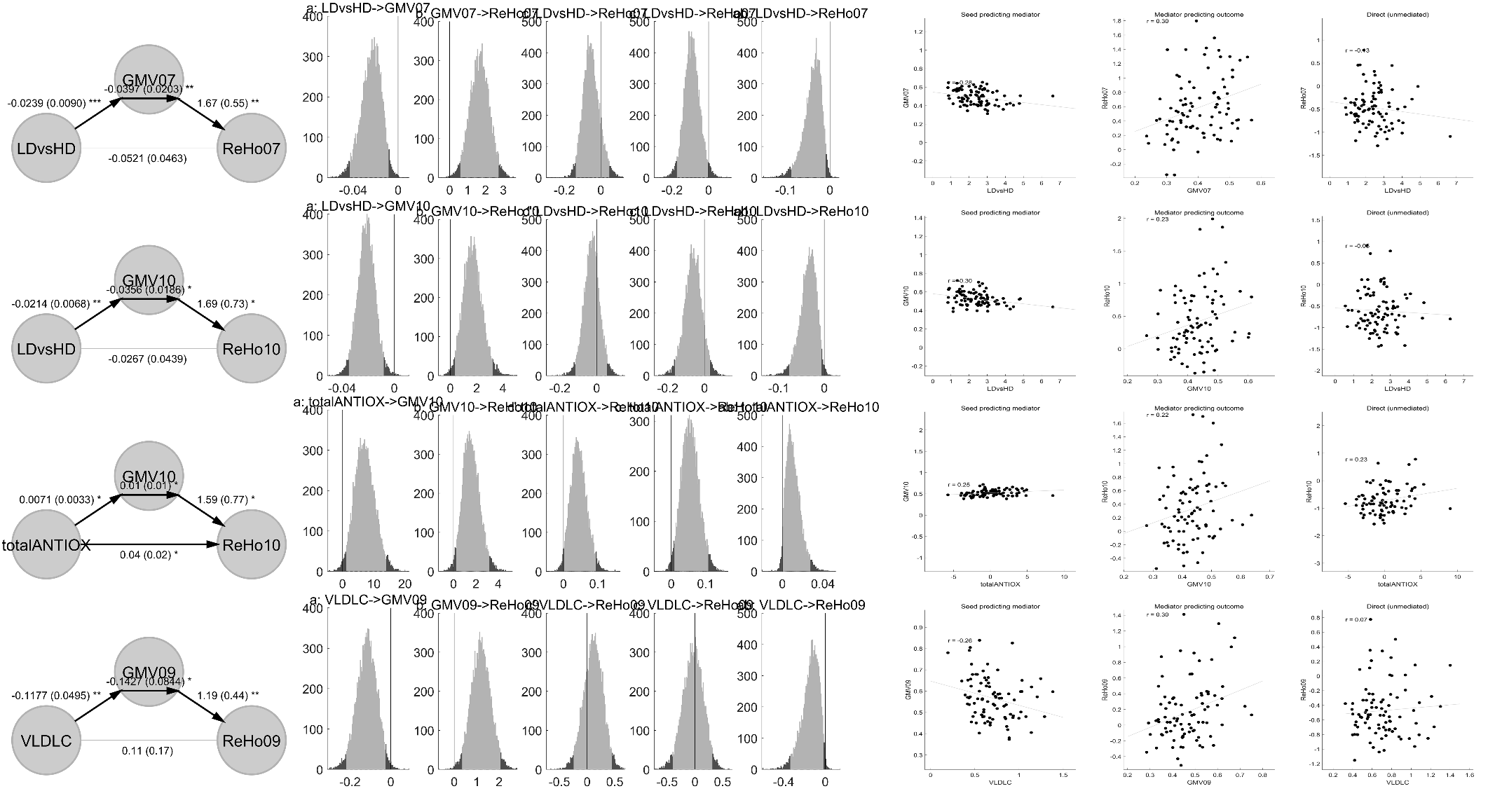

**ESF, Figure 3** Mediation analyses linking peripheral biomarkers to regional gray matter volume (GMV) and fractional amplitude of ReHo. Each row presents one significant mediation model in which a peripheral biomarker was entered as the predictor (X), regional GMV as the mediator (M), and regional ReHo as the outcome (Y). Left panels show the mediation path diagrams, including the X→M (path a), M→Y controlling for X (path b), and direct X→Y effect controlling for M (path c′). Values are regression coefficients with standard errors in parentheses; asterisks indicate statistical significance (P < 0.05, *P < 0.01, **P < 0.001). Middle panels show the bootstrap distributions of the mediation-path coefficients, including the indirect effect (ab). Right panels show the corresponding partial-regression scatterplots for the predictor–mediator association, mediator–outcome association, and direct predictor–outcome association. All mediation models were adjusted for age, sex, and body mass index and were evaluated using 10,000 bootstrap samples. GMV, gray matter volume; ReHo: regional homogeneity; HDL-C, high-density lipoprotein cholesterol; LDvsHD, low-density lipoprotein cholesterol/high-density lipoprotein cholesterol ratio; totalANTIOX, the NAPR index.
